# Feasibility and validity of using self-collected capillary blood using Tasso+ for measuring Alzheimer’s Disease plasma-based biomarkers among underrepresented populations

**DOI:** 10.64898/2026.02.02.26345372

**Authors:** Amy A Schultz, Adam J Paulsen, Aaron Fredricks, David T Plante, Paul E Peppard, Rachael Wilson

## Abstract

**Background:** Blood-based biomarkers offer a scalable alternative to cerebrospinal fluid and PET imaging for Alzheimer’s disease (AD) detection, yet traditional venipuncture limits participation among rural and socioeconomically disadvantaged populations. Self-collection using the Tasso+ capillary device could reduce access barriers, but its feasibility and validity for AD plasma biomarkers remain uncertain, particularly with real-world delays prior to processing.

**Methods:** Adults aged 45–90 years from the Wisconsin SHOW cohort who were underrepresented in AD research (Black or Hispanic race/ethnicity, rural residence, or <bachelor’s degree) were recruited (n=28). At community “pop-up” clinics participants completed: (1) self-collection of capillary blood via Tasso+; (2) experience surveys; (3) Montreal Cognitive Assessment; and (4) standard venipuncture. To simulate home-based collection and mail return, Tasso+ samples were held at room temperature for 24 hours before centrifugation, whereas venous samples were processed within 30 minutes. Plasma Aβ40, Aβ42, Aβ42/40, GFAP, NfL, and pTau217 were measured on the Quanterix Simoa platform. Between-method agreement was evaluated using Pearson/Spearman correlations, Lin’s concordance correlation coefficients (CCC), Bland–Altman analyses, and relative bias. Predictors of percent difference were explored with univariate regression.

**Results:** Tasso+ collection was successful for 96% of participants; 64% rated it very easy and 86% reported comfort/no pain, yet 57% preferred future venipuncture—particularly Black, lower-income, and lower-education participants. Agreement varied markedly by biomarker. GFAP and NfL demonstrated excellent concordance (CCC 0.97–0.98) with minimal bias (–6% to –8%). Aβ40 and Aβ42 showed modest correlations (r=0.40–0.47) and substantial underestimation (–60% to –70%). Aβ42/40 and pTau217 exhibited poor correlation and extreme positive bias for pTau217 (∼+2600%). Hemolysis was more frequent in Tasso+ samples and contributed to disagreement for several markers; processing lag and sample volume were not strong predictors.

**Conclusions:** Remote capillary self-collection with a 24-hour delay is suitable for measuring GFAP and NfL but not currently reliable for Aβ or pTau217 without improved handling (e.g., temperature control, hemolysis reduction). Although user experience was favorable, trust and logistical concerns limited preference among underrepresented groups. Community-informed strategies and optimized pre-analytics are essential before deploying Tasso+ in large AD studies.

## Introduction

A major challenge facing Alzheimer’s Disease (AD) research is the heterogeneity of those at risk for AD and the heterogeneity of underlying pathologies and mechanisms.^1,2^ Among those aged 65+ years of age, AD risk is highest among Black (19.3%) and Hispanic (16.7%) individuals when compared to non-Hispanic White individuals (7.4%).^1^ Furthermore, risk-adjusted AD diagnostic incidence is higher in rural versus urban counties, despite a lower prevalence, suggesting underdiagnosed AD in rural communities.^3^ The variability seen among those with AD may result from multiple factors, including co-morbidities, socioeconomic factors, biological risk factors (genetics and epigenetics), environmental and behavioral factors, and/or cognitive resilience or reserve.^4^ Yet, the populations most affected by AD – rural, low socio-economic status, low educational attainment, and ethnoculturally diverse groups – are the same populations underrepresented in AD research studies.^4,5^ Aging research requires greater representation to disentangle the socio-cultural, behavioral, and environmental factors across the life course that are associated with aging and may contribute to AD disparities.^3^

Earlier and easier detection of neurodegenerative changes would aid in identifying modifiable risk factors, prevention strategies, and those in need of intervention. AD diagnosis has widely relied on amyloid-PET imaging and/or the measure of amyloid-beta (Aβ) peptides, tau, and phosphorylated-tau (pTau) concentrations within cerebrospinal fluid (CSF).^6^ These techniques require a clinic visit at specific medical centers, are expensive, and invasive – a major contributor to underdiagnosis and a lack the of representation from the populations most at risk of AD in aging research. Study visit burden, distance to major hospital clinics, and lack of transportation or insurance are major contributors to the underlying underrepresentation.^6,7^

To address the lack of diversity in AD research, recent studies have shown feasibility of conducting venipuncture blood collection at remote sites for plasma biomarker testing as a more accessible approach compared to the standard CSF testing and PET imaging used for AD diagnosis.^6,7^ Blood-based biomarkers have been shown to be associated with CSF or PET biomarkers and can effectively differentiate AD from other neurodegenerative diseases.^4^ Yet, recent advancements in single molecule array, ultrasensitive technology, enable detection of blood proteins at sub-femtomolar (very low) concentrations.^8^ This opens the door for the possibility of using self-administered, capillary blood collection to measure blood biomarkers for AD.

Self-administered blood collection has the potential to increase access to diagnostic testing and to expand participation in aging research, especially among those with limited mobility, an aversion to healthcare settings or needles, or burdened by the travel.^9^ Collection kits, such as the Tasso+ (Tasso, Inc., Seattle, WA), are designed to be conducted in one’s home and mailed to a laboratory for analysis. Only a few studies have compared venous blood from venipuncture blood draw to capillary blood from a Tasso device (i.e., examining liver chemistry,^10^ infectious disease or protein/inflammatory markers,^11^ lipids and proteomics),^12,13^ but they report high correlations and concordance. Participant feedback was also favorable of the Tasso+ experience, noting lancet puncture on the arm to be less painful than finger lancet or needle puncture from venous blood draw.^10,14^ To our knowledge, only one study to-date has used Tasso+ to measure Neurofilament light chain (NfL) and glial fibrillary acidic protein (GFAP), but on only n=1 due to serious aversion to needles.^15^ While recent studies have tested blood biomarkers’ utility in clinical practice,^16^ limited to no testing of the blood biomarkers has been conducted in a non-clinical population or underrepresented population using self-administered blood collection.

Thus, the following pilot aims to (1) assess the feasibility of self-administered capillary blood collection using Tasso+ among underrepresented populations– *are people willing and interested in using Tasso+?* – and (2) examine the validity of measuring AD blood biomarkers from capillary blood using Tasso+ compared with venipuncture. The goal of the study was to mimic conditions of a large, prospective cohort study where self-administered blood collection kits are mailed to adults and returned within 24 hours to a laboratory for analysis.

## Materials and Methods

### Study Population

The study sample was recruited from the Survey of the Health of Wisconsin (SHOW) cohort. SHOW is the only statewide cohort modeled after the CDC’s NHANES study, with baseline survey data and biospecimen collected on over 5,842 adults from 2008-2019.^17^ Participants include residents from rural and urban areas across the state. In 2019, SHOW dedicated recruitment efforts to increasing representation from Black and Hispanic communities in Milwaukee, thereby increasing the racial and ethnic diversity of the cohort.^17^ Participants consent to be contacted for future ancillary studies. SHOW is a general health survey and participants were not specifically recruited for AD research.

### Eligibility Criteria

To determine whether Tasso+ is feasible and valid to use among populations most affected by AD, SHOW adults needed to meet the following criteria:

1. Current age 45-90 years
2. Highest educational attainment less than a bachelor’s degree
3. Self-reported one or more of the following:
  a. Black or African American race,
  b. Hispanic or Latino ethnicity,
  c. reside in rural area
4. Reside in Dane, Milwaukee, or Waushara counties in Wisconsin

Waushara County was selected as it has SHOW participants in rural areas >80 miles from any of the large metropolitan centers in the state – Milwaukee, Madison, and Green Bay. Milwaukee and Dane counties were selected because they have higher concentrations of SHOW participants who identify as Black and/or Hispanic or Latino. Adults were considered to reside in a rural area if their residential address was not within an urban area, as defined by the U.S. Census (urban areas have at least 5,000 people or 2,000 housing units).^18^

### Recruitment

To avoid recruiting more individuals than necessary to meet the recruitment goal of 25-30 participants, batches of n=20 adults were randomly selected by county and sent recruitment invitations. Additional batches of n=20 were recruited biweekly until 10-12 appointments were scheduled in each county; a total of 100 invites went out. Selected adults were sent up to two mailed invitations, spaced one week apart, and up to three emailed invitations (if email address was available), spaced 1 week apart. The invitation contained a website address and QR code where interested adults completed a screener (verifying eligibility criteria above) and scheduled their appointment location, day, and time. A phone number was also provided for participants to complete the screener and schedule their appointment over the phone with a study staff member. All recruitment, scheduling, and study appointment data were collected in Research Electronic Data Capture (REDcap), a secure, HIPAA-compliant web application for research data collection and management of patient health information (PHI).^19^ The REDcap project is administered and managed by the UW-Madison Institute for Clinical & Translational Research (ICTR).

### Study Appointment Overview

All study procedures were approved by the Health Sciences Institutional Review Board at the University of Wisconsin-Madison. Participant appointments were held at a pop-up clinic set up in a community site location – one in each county. The sites were chosen because they are trusted community locations used by SHOW in prior years.

Appointments were conducted by an experienced study coordinator trained in phlebotomy and survey interviewing methods. The appointment took 30-45 minutes and consisted of the following elements: informed consent, self-collection of capillary blood using a Tasso+ device, an interview survey about their experience, the Montreal Cognitive Assessment (MoCA),^20^ a venous blood draw performed by the staff member, and a final interview survey evaluating their preference between using Tasso+ or having a venipuncture blood draw (See Figure 1 for a diagram overview). Participants provided written informed consent prior to study activities and received $75 for completing the study.

**Figure 1.**
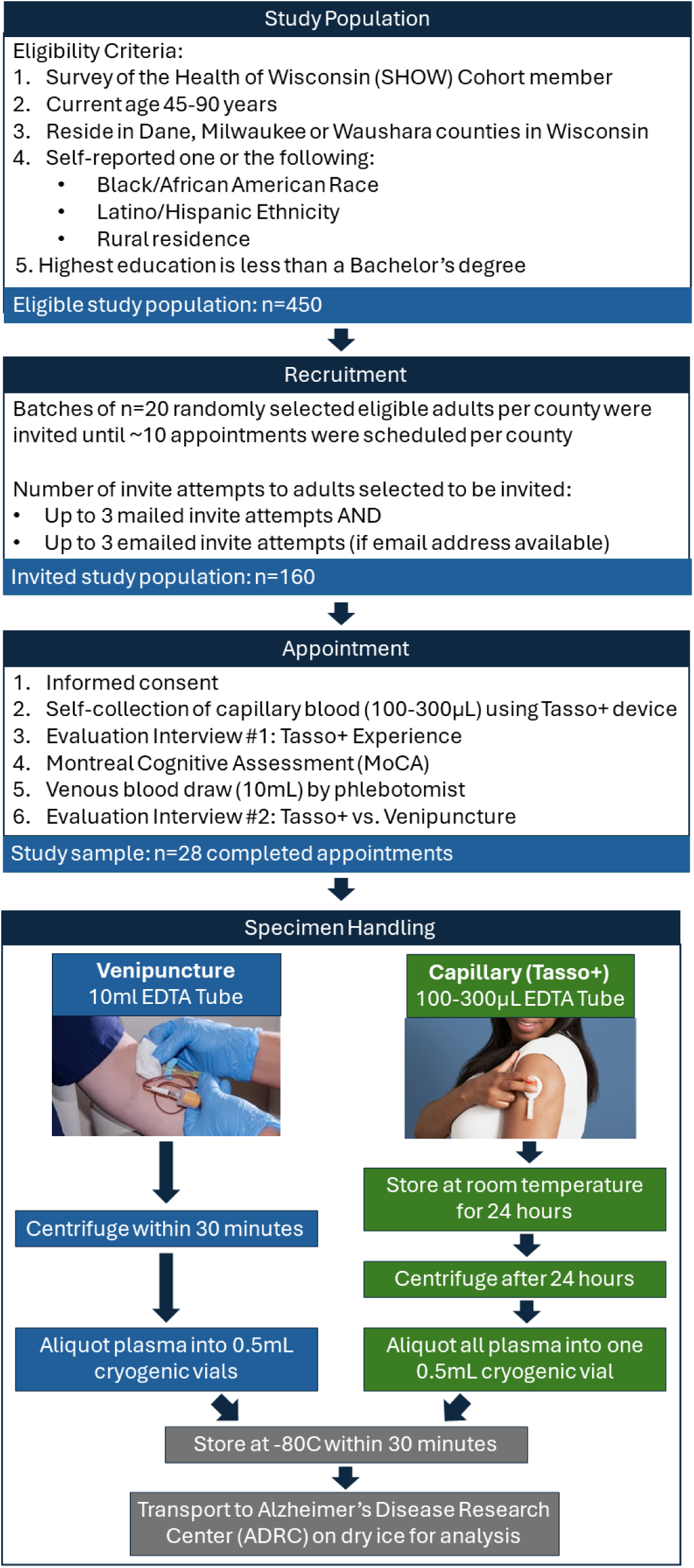
Overview of the study design.

### Blood Collection

The Tasso+ device with the EDTA tube collection kit (Tasso, Inc., Seattle, WA) was used for self-collection. Tasso+ is a U.S. Food and Drug Administration Class II Lancet 510(k)-cleared device. Participants were asked to read paper instructions and watch video instructions prior to conducting the self-collection (See Supplementary materials for the paper and video instructions provided by the company). The study coordinator answered any questions prior to the participant starting the self-collection. The participant collected up to 300 µL of blood using the Tasso+ device, which adheres to the upper arm and pokes the arm (like a finger lancet prick). The device deposits capillary blood into an attached, mini EDTA tube. Participants were given the option to use either or both instructions (paper and video) while performing the self-collection themselves. The study coordinator assisted only if needed throughout the process, and any assistance was recorded. If participants failed at collecting blood, up to two additional attempts were allowed if they were willing. After the participant completed the collection steps, they removed the mini EDTA tube and inverted it several times (per the instructions). Finally, they were instructed to hand it to the study coordinator for the remaining specimen handling.

A venipuncture blood draw was performed on the opposite arm by the study coordinator into a 10mL EDTA tube. The study coordinator recorded the last time the participant ate and the time of both blood collections.

### Survey Interviews

#### Demographic and health survey

Participants were asked to confirm their residential address, date of birth, sex, race, ethnicity, highest educational attainment, annual household income and the number of people supported on the household income. Participant responses from their last SHOW survey were populated prior to their appointment. Any updates to their information were recorded in REDcap.

#### Tasso+ Experience Evaluation

Following the self-collected blood, the study coordinator administered a 7-item survey interview about their experience using Tasso+. The items asked how useful the instructions were in guiding them through the process, what the most confusing step was, how difficult or easy it was, what the most difficult part of the collection process was, whether they felt any pain, and how willing and confident they would be to use the device on their own in the future. Two items were open-ended, and five items had Likert scale response options. See supplementary materials to view the survey.

#### Montreal Cognitive Assessment (MoCA)

In between the self-collection of capillary blood and the venipuncture draw, the study coordinator administered the Montreal Cognitive Assessment (MoCA). The MoCA is a widely used screening tool for mild cognitive impairment that evaluates attention, executive function, memory, language, visuospatial abilities, and orientation. Scores range from 0 to 30 and were categorized for descriptive characterization of the study sample as ≥ 26, 18-25, and 10-17, reflecting unimpaired cognition, mild impairment, and more substantial impairment, respectively.^20^

#### Tasso+ vs. Venipuncture Evaluation

The final survey interview was administered after the venipuncture blood draw. The 3-item evaluation asked participants to compare the Tasso+ to the venipuncture: which method was more painful, which method would they prefer in the future, and why did they prefer their chosen method. They could also provide additional miscellaneous feedback.

All interview responses and MoCA scores were recorded in REDcap. See supplementary materials to view the final evaluation survey.

### Specimen Handling

The 10mL EDTA tube from venipuncture was centrifuged within 30 minutes of collection. The tube was inverted 10-12 times, centrifuged for 10 minutes at 2000g at room temperature, and aliquoted into 0.5mL cryogenic vials. The plasma specimen was stored in a −80C freezer at UW-Madison until delivery to the lab for analysis. While specimens were immediately put in −80C freezer following Dane County site appointments, following Waushara and Milwaukee County site appointments, specimens were immediately placed on dry ice for transport to UW-Madison and stored in −80C within 8 hours of collection.

To mimic self-collection of blood using Tasso+ via a mailed kit that is returned via mail overnight to the lab, the mini-EDTA tube of capillary blood was stored at room temperature for 24 hours prior to centrifuging. After 24 hours, the mini-EDTA tube was processed following the procedures outlined above for the venipuncture collected blood. The study coordinator recorded the time of centrifuging and whether any hemolysis was present after aliquoting.

### Laboratory Analysis

All plasma specimens were delivered on dry ice to the Alzheimer’s Disease Research Core Biomarker Laboratory at the University of Wisconsin–Madison. Samples were analyzed on HD-X (Quanterix, Billerica, MA) platform using the Simoa® ALZpath p-Tau217 Advantage PLUS assay (Quanterix PN 104465, LN 504182, Exp date 23-Aug-2025) and the Simoa® Neurology 4-Plex E PLUS Kit (Quanterix PN, LN, Exp date) to quantify Aβ40, Aβ42, GFAP, and NfL. Given the limited volume for samples collected via Tasso+, p-Tau217 measurements were prioritized over Neurology 4-plex E (ie. if there was not enough volume for both assays, only p-Tau217 was assayed). Plasma p-tau217 has recently become a leading blood-based marker of AD pathophysiology, exhibiting high congruence with CSF and with brain amyloid and tau PET signal across the AD spectrum.^21,22^ Samples were analyzed in duplicate when adequate volume was available, and prepared and analyzed according to manufacturer instructions. Both manufacturer controls and pooled plasma QC controls prepared in-house were analyzed in duplicate on the same plate to assess accuracy and precision. On the day of analysis, samples were removed from the −80°C freezer and thawed to room temperature. For pTau217, approximately 100 μL of each sample was transferred to a 96-well plate for analysis. The remaining volume, ranging from 30-90 μL, was transferred to a separate 96-well plate for analysis with the Neurology 4-plex E kit.

### Data and Statistical Analysis

#### Variable treatment

SAS (version 9.4) and R (version 4.5.2) were used for conducting data cleaning and data analysis, respectively. Current age was categorized when describing the study sample (48-64, 65-74, and 75-90 years) but left as continuous (10-year increments) in univariate regression. U.S. Federal Poverty Level (FPL) - below 100% FPL, or at or above 100% FPL – was determined via the U.S. Department of Health and Human Services’ annual poverty guidelines from self-reported household income and the number of people supported.^23^ Highest educational attainment was examined as 3-categories: 2^nd^-10^th^ grade, 11-12^th^ (no diploma), and ≥ HS diploma/GED and 2-categories: <HS diploma/GED vs. ≥ HS diploma/GED. While all participants had less than a bachelor’s degree, some had Associates or vocational/trades degrees or attended some college classes. Body mass index (BMI, in kg/m^2^) was calculated from participants’ last SHOW survey using measured weight and height. BMI was examined as < 30 kg/m^2^ (non-obese) vs. ≥ 30 kg/m^2^ (obese) based on the CDC’s BMI Categories.^24^

Residential addresses were geocoded using CENTRUS and linked to U.S census county and census block group data. Individuals were considered to reside in a moderately to highly deprived area if their residence was in a block group that ranks nationally in the 6th-10th decile based on the Area Deprivation Index.^25^

#### Tasso+ Feasibility

Frequencies and percents of survey responses from the evaluation interviews are presented. Chi-square test was used to determine whether blood draw preferences (Tasso+ vs. venipuncture) statistically differed by demographic, socioeconomic, geographic, and other health characteristics.

#### Tasso+ validity for measuring ADRD plasma biomarkers

##### Summary Statistics

We conducted all analyses using biomarker concentrations measured in plasma collected via the Tasso+ capillary device or venipuncture. When duplicate runs were successful, the average of the two concentrations was used. Singlet concentrations were reported when duplicates were not available. To characterize biomarker distributions for each method separately, we calculated summary statistics including the minimum, maximum, mean, standard deviation (SD), median, and interquartile range (IQR). The coefficient of variation (CV) was defined as the ratio of the SD to the mean and expressed as a percentage: *CV* = (*SD*/*mean*) × 100. CV quantifies relative variability and allows comparison of dispersion across biomarkers with different concentration ranges.

##### Between-method agreement

To evaluate whether Tasso+ concentrations can be used interchangeably with venipuncture—we restricted analyses to participants with valid paired plasma samples for each biomarker. We calculated Pearson and Spearman correlation coefficients to examine linear and rank-order relationships. Agreement was assessed using Lin’s concordance correlation coefficient (CCC) and Bland–Altman methods. Lin’s CCC evaluates how well paired measurements fall on the 45° line of equality, incorporating both correlation and mean bias. Relative bias was defined as

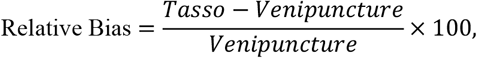

and mean difference in biomarker concentration was computed as *Tasso* − *Venipuncture*. Ninety-five percent confidence intervals (CIs) for bias and differences were obtained using nonparametric bootstrap resampling, and Bland–Altman plots display overall bias and limits of agreement.

##### Predictors of percent (%) mean difference

We modeled the percentage difference as the outcome in univariate linear regression models calculated as:

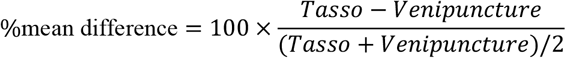

Predictors examined included demographic characteristics, cognitive function from MoCA, subjective Tasso+ experience, hemolysis status (yes vs. no), and sample volume. To examine whether plasma sample handling and processing affected the difference in concentrations from Tasso+ and venipuncture, the number of laboratory assay runs (singlet or duplicate) were examined, and so was the time lag from collection to centrifuging and the time since the participant last ate. The 4-to-5-point Likert responses to Tasso+ experience evaluation were collapsed into 2-categories for analysis as follows: Tasso difficulty was defined as “Very easy” or combining remaining responses into “Difficult”; Tasso comfort was defined as “Comfort or no pain” or combining remaining responses into “Discomfort or pain”; Tasso willingness was defined as “Very willing” or combining remaining responses into “Not willing”; and Tasso confidence was defined as “Very Confident” or combining remaining responses into “Not Confident”. Models were fit separately for each biomarker, without adjustment for multiple comparisons, and robust standard errors (HC2) were used to account for small sample sizes.

##### Within-method repeatability

In addition to the main analysis, we evaluated the within-method repeatability using only those participants with duplicate laboratory runs. For each participant and biomarker, we computed the mean concentration across replicates and quantified within-person variation using the replicate SD and CV. These metrics assess analytic precision (repeatability of the assay under identical conditions) rather than accuracy. Because venipuncture samples universally had duplicate runs and Tasso+ samples varied (singlet vs. duplicate), we present within-method summary variability separately for each method.

## Results

### Study sample characteristics

A total of 28 adults were enrolled across three Wisconsin counties, with one-third from rural areas (Table 1; Figure 1). Participants ranged in age from 48–90 years (mean = 69.8, SD = 9.2). Slightly over half were women (51.9%), 42.9% self-identified as Black or African American, and 7.1% identified as Hispanic or Latino. Most participants (71.4%) were obese (BMI ≥ 30), 39.3% lived below the federal poverty level, 64.3% lived in areas with high neighborhood deprivation, and 57.1% had not completed a high school degree or GED. Cognitive performance was varied: with 42.9% scored ≥26 on the MoCA (no impairment), 42.9% demonstrated mild impairment (18–25), and 7.1% scored 10–17.

**Table 1.** Study sample (n=28) descriptives by their preference for future blood collection: self-collection via Tasso+ at home vs. venipuncture via typical blood draw at a clinic.

|  |  | Total<br>(n=28) |  | Preference<br>Tasso+ Self Clinic Blood<br>Collection Draw<br>(n=12) (n=16) |  |  |  |  |
| --- | --- | --- | --- | --- | --- | --- | --- | --- |
| Selected Characteristics |  | N | % | N | Col % | N | Col % | P-value |
| Age | Mean (SD) | 69.8 | 9.2 | 73 | 6.6 | 67.4 | 10.3 |  |
| Age |  |  |  |  |  |  |  | 0.31 |
|  | 48-64 | 9 | 32.1 | 2 | 16.7 | 7 | 43.8 |  |
|  | 65-74 | 10 | 35.7 | 5 | 41.7 | 5 | 31.3 |  |
|  | 75 or older | 9 | 32.1 | 5 | 41.7 | 4 | 25.0 |  |
| Sex |  |  |  |  |  |  |  | 0.86 |
|  | Male | 13 | 46.4 | 6 | 50.0 | 7 | 43.7 |  |
|  | Female | 15 | 53.6 | 6 | 50.0 | 9 | 56.3 |  |
| Race |  |  |  |  |  |  |  | 0.002 |
|  | White | 16 | 57.1 | 11 | 91.7 | 5 | 31.3 |  |
|  | Black or African American | 12 | 42.9 | 1 | 8.3 | 11 | 68.8 |  |
| Ethnicity: Hispanic or Latino |  |  |  |  |  |  |  | 0.21 |
|  | Yes | 2 | 7.1 | 0 | 0.0 | 2 | 12.5 |  |
|  | No | 26 | 92.9 | 12 | 100.0 | 14 | 87.5 |  |
| Combined Annual Household Income |  |  |  |  |  |  |  | 0.0013 |
| | < \$20,000 | 9 | 32.1 | 0 | 0.0 | 9 | 56.3 | |
| | \$20,000 to \$59,000 | 11 | 39.3 | 5 | 41.7 | 6 | 37.5 | |
| | ≥ \$60,000 | 8 | 28.6 | 7 | 58.3 | 1 | 6.3 | |
| Federal Poverty Level (FPL)<sup>a</sup> |  |  |  |  |  |  |  | 0.0007 |
|  | Below 100% FPL | 15 | 53.6 | 1 | 8.3 | 11 | 68.8 |  |
|  | At or above 100% FPL | 12 | 42.9 | 11 | 91.7 | 4 | 25.0 |  |
|  | Missing | 1 | 3.6 |  |  | 1 | 6.3 |  |
| Highest Educational Attainment |  |  |  |  |  |  |  | 0.039 |
|  | 2nd-10th grade | 9 | 32.1 | 1 | 8.3 | 8 | 50.0 |  |
|  | 11th-12th grade (no diploma) | 7 | 25.0 | 3 | 25.0 | 4 | 25.0 |  |
|  | HS diploma/GED or more | 12 | 42.9 | 8 | 66.7 | 4 | 25.0 |  |
| Wisconsin County |  |  |  |  |  |  |  | 0.003 |
|  | Dane | 8 | 28.6 | 7 | 58.3 | 1 | 6.3 |  |
|  | Milwaukee | 11 | 39.3 | 1 | 8.3 | 10 | 62.5 |  |
|  | Waushara | 9 | 32.1 | 4 | 33.3 | 5 | 31.3 |  |
| Census Urban / Rural<sup>b</sup> |  |  |  |  |  |  |  | 0.91 |
|  | Urban | 19 | 67.9 | 8 | 66.7 | 11 | 68.8 |  |
|  | Rural | 9 | 32.1 | 4 | 33.3 | 5 | 31.3 |  |
| Area Deprivation Index (ADI)<sup>c</sup> |  |  |  |  |  |  |  | 0.0045 |
|  | Low to moderate deprivation 1-5 | 10 | 35.7 | 8 | 66.7 | 2 | 12.5 |  |
|  | Moderation to High deprivation 6-10 | 18 | 64.3 | 4 | 33.3 | 14 | 87.5 |  |
| MoCA Total Score |  |  |  |  |  |  |  | 0.31 |
|  | <10 | 0 | 0.0 | 0 | 0.0 | 0 | 0.0 |  |
|  | 10-17 | 2 | 7.1 | 2 | 16.7 | 7 | 43.8 |  |
|  | 18-25 | 12 | 42.9 | 5 | 41.7 | 5 | 31.3 |  |
|  | ≥26 | 12 | 42.9 | 5 | 41.7 | 4 | 25.0 |  |
|  | Missing | 2 | 7.1 |  |  |  |  |  |
| Body Mass Index (BMI)<sup>d</sup> |  |  |  |  |  |  |  | 0.69 |
|  | <30 | 8 | 28.6 | 4 | 33.3 | 4 | 25.0 |  |
|  | ≥30 | 20 | 71.4 | 8 | 66.7 | 12 | 75.0 |  |

Abbreviations: Col – column; HS – High School; GED – General Education Development; ER – Emergency Room; MoCA – Montreal Cognitive Assessment.

### User Experience with Tasso+

Most participants reported a highly favorable experience using Tasso+ (Figure 2). Two-thirds (64.3%) rated the device as very easy to use, 85.7% experienced comfort or no pain, and 75% were very willing to use Tasso+ again. No participants reported moderate or severe discomfort, and none expressed unwillingness to use the device in the future.

**Figure 2.**
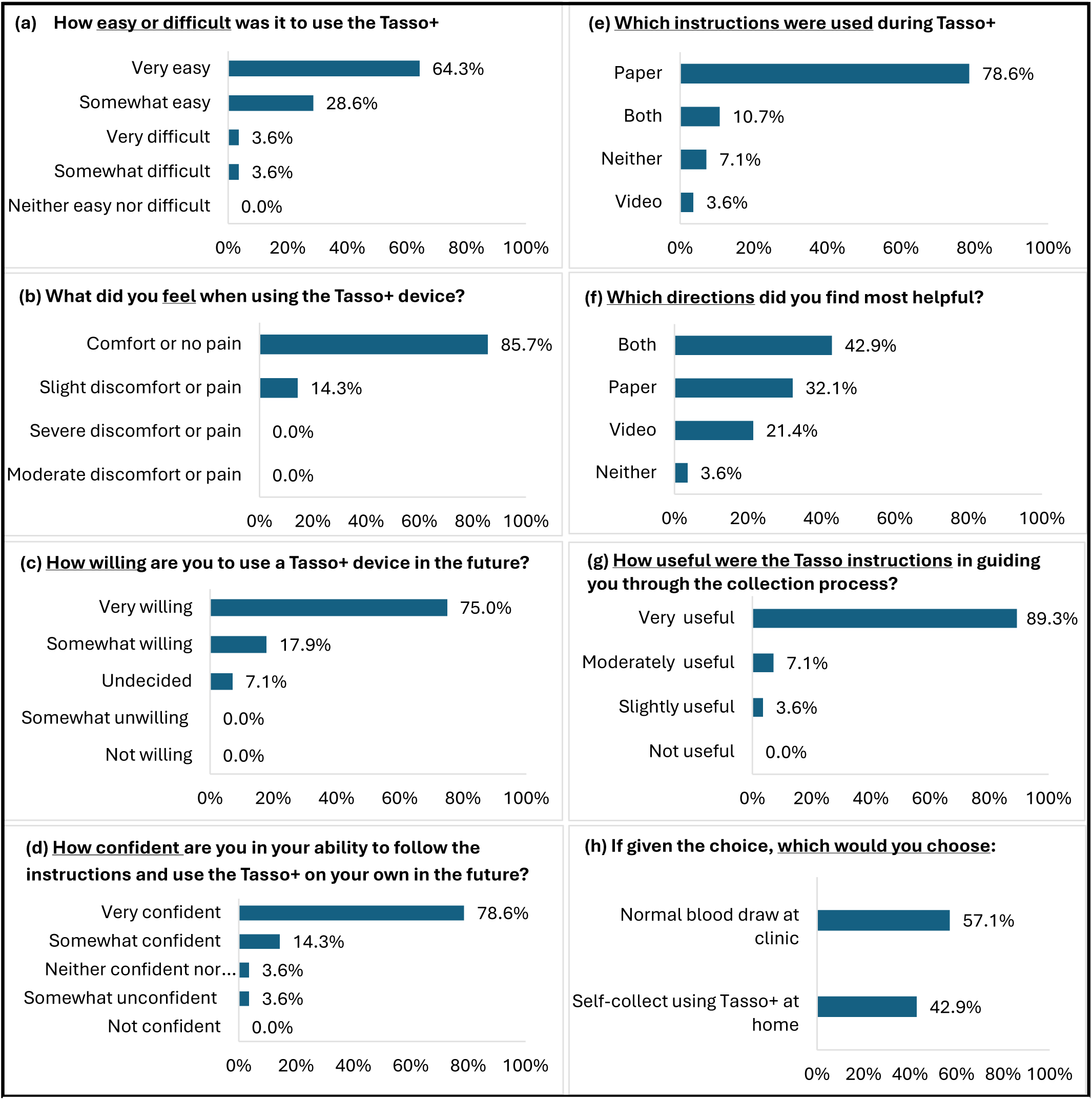
Participant responses (n=28) to the Tasso+ experience evaluation and the Tasso+ vs. Venipuncture evaluation. Both were administered via interview at the study appointment, immediately following Tasso+ blood collection and venipuncture blood collection.

Paper instructions were used most commonly (78.6%), while 10.7% used both video and paper, 3.6% used video alone, and 7.1% used neither. Most participants rated both formats as helpful, although qualitative feedback (Table S1) highlighted challenges and areas for improvement. Most notable were difficulties verifying how hard to push the button (did not know to anticipate a “click” sound), knowing whether blood was flowing adequately or how long to wait for blood to appear, and performing physical steps such as aligning or twisting components, particularly for participants with neuropathy or vision impairment. A few participants also mentioned the complexity of the multi-step instructions.

### Future Preference: Tasso+ vs. Venipuncture

Despite positive experiences, only 43% ultimately preferred Tasso+ for future blood draws, compared with 57% who preferred venipuncture (Table S2). Those who preferred Tasso+ emphasized that the device was easier or more convenient than traveling to a clinic site, took less time, and could be completed at home, because as one participant stated, they “live in the middle of nowhere”. Some participants also viewed Tasso+ as a particularly attractive option for when frequent blood testing is needed. Those preferring venipuncture emphasized discomfort with the responsibility and/or difficulty of collecting and mailing their own sample; others had concerns about mailing reliability (i.e. sample getting lost, stolen, or quality degrading).

Participant characteristics differed by future blood collection preference (Table 1). Following both Tasso+ and venipuncture methods, those who preferred using Tasso+ were more likely to be White, have higher income, live above the federal poverty level, hold a high-school/GED degree, and reside in low-deprivation areas. Conversely, participants from underrepresented populations in AD research–particularly Black participants, those with the lowest educational attainment (2^nd^-10^th^ grade), and those living in high deprivation neighborhoods–were much more likely to prefer a venipuncture blood draw at clinic over using Tasso+ at home. Differences were not seen by rural-urban status.

### Blood Collection and Biomarker Analysis

Venipuncture was successfully completed for 26 of 28 participants (93%), while Tasso+ samples were obtained from 27 of 28 participants (96%) (Table S2). Most Tasso+ collections required a single attempt, although four participants required multiple attempts; 25 were self-collections (89%), 2 needed assistance due to visual or motor limitations, and one was unsuccessful.

Hemolysis was more common in Tasso+ samples than venipuncture samples: moderate-to-full hemolysis occurred in seven Tasso+ samples compared with one venipuncture sample (Table S3). Tasso+ plasma volumes varied widely (Figure S1), with most participants yielding 150–300 μL, and only a small number producing volumes insufficient for all planned assays. Data completeness for Tasso+ reflected this variation: usable concentrations were obtained for 23 participants for pTau217 (prioritized assay), 17 for Aβ40, 16 for Aβ42 and Aβ42/40, and 18 for GFAP and NfL, compared with 26 venipuncture samples for all biomarkers. Additionally, duplicate analyses were not possible for all the Tasso+ samples (Table S3).

Summary statistics (Table 2; Table S5) indicated large between-person variability for all biomarkers. Tasso+ and venipuncture showed similar SDs for GFAP, NfL, and Aβ measures, although coefficients of variation were often higher for Tasso+, reflecting greater relative variability.

**Table 2.** Summary statistics on the distribution and variability of each biomarker (in pg/mL) for Tasso+ vs. venipuncture collection method.

| Biomarker | Tasso+ Capillary |  |  |  |  |  | Venipuncture |  |  |  |  |  |
| --- | --- | --- | --- | --- | --- | --- | --- | --- | --- | --- | --- | --- |
|  | N | Min | Max | Mean (SD) | CV | Median (IQR) | N | Min | Max | Mean (SD) | CV | Median (IQR) |
| Aβ40 | 17 | 1.99 | 109.15 | 57.70 (29.4) | 50.93 | 61.3 (24.0) | 26 | 101.50 | 205.70 | 139.40 (25.3) | 18.11 | 130.50 (34.4) |
| Aβ42 | 16 | 0.32 | 3.91 | 2.53 (01.1) | 41.68 | 2.73 (01.4) | 26 | 5.13 | 11.47 | 7.93 (01.6) | 19.69 | 7.60 (02.1) |
| Aβ42/40 | 16 | 0.03 | 0.10 | 0.05 (0.02) | 37.50 | 0.04 (0.01) | 26 | 0.05 | 0.07 | 0.06 (0.01) | 9.18 | 0.06 (0.01) |
| GFAP | 18 | 35.68 | 252.71 | 91.94 (49.8) | 54.18 | 87.02 (47.7) | 26 | 38.28 | 251.63 | 98.32 (51.5) | 52.42 | 89.90 (42.8) |
| NfL | 18 | 6.50 | 51.05 | 18.43 (10.4) | 56.59 | 16.39 (07.0) | 26 | 6.13 | 50.21 | 19.00 (10.1) | 53.36 | 18.23 (09.2) |
| pTau217 | 23 | 1.71 | 18.42 | 6.98 (03.9) | 55.93 | 6.34 (04.2) | 26 | 0.14 | 0.99 | 0.34 (00.2) | 63.52 | 0.29 (00.2) |

### Biomarker Descriptives

Violin plots of raw concentrations (Figure 3) illustrate that, at the group level, Tasso+ concentrations were *much higher* than venipuncture concentrations for pTau217, *lower* for Aβ40 and Aβ42, *roughly similar* for Aβ42/40, and *very similar* for GFAP and NfL–consistent with corresponding summary statistics (Table 2; Table S5) which indicated large between-person variability (SD and CV) for all biomarkers except GFAP and NfL. Tasso+ and venipuncture showed similar SDs for GFAP, NfL, and Aβ measures, although CVs were often higher for Tasso+, reflecting greater relative variability. In contrast, while CV was comparable for pTau217, the mean and SD were much greater for Tasso+. Concentrations were lower across all biomarkers among singlet-run samples compared with duplicate-run samples for Tasso+ collected plasma (Table S5).

**Figure 3.**
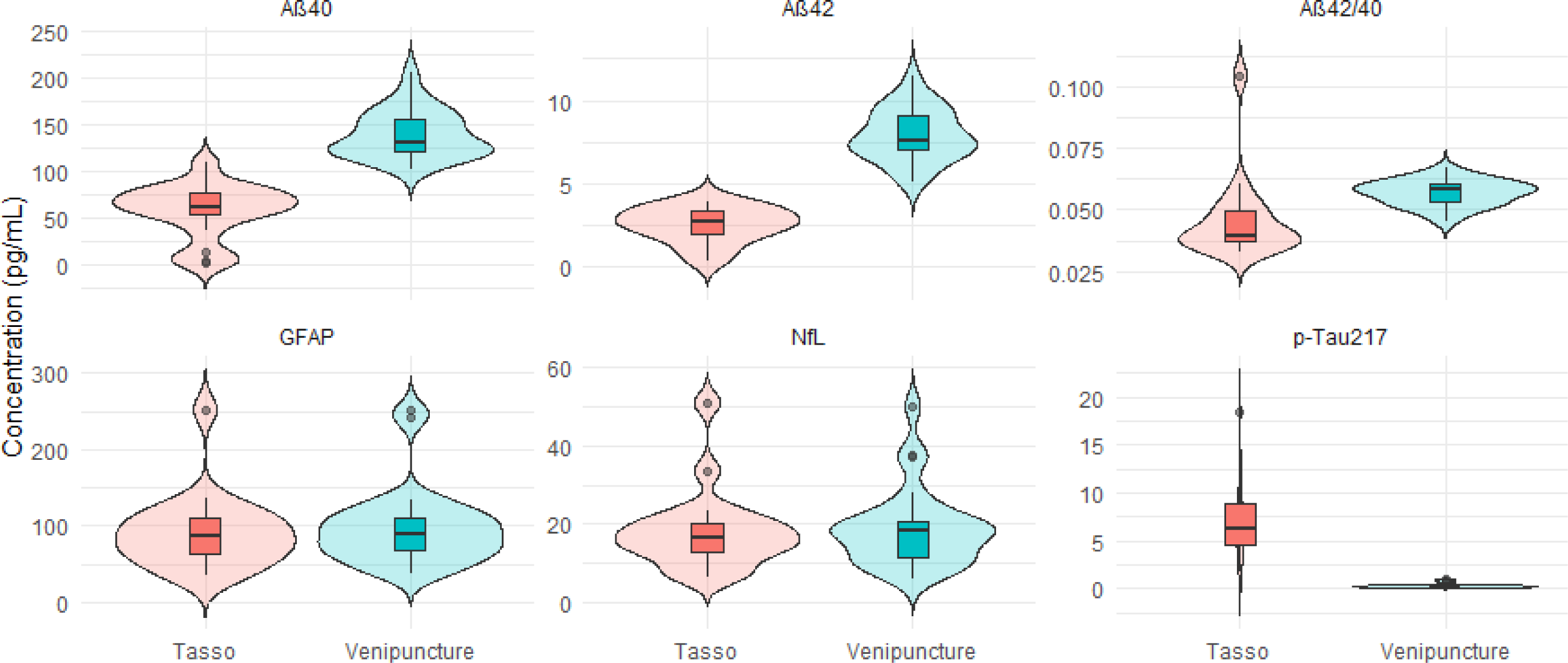
Violin plots depicting the distribution of concentrations (pg/mL) measured for each of the six ADRD biomarkers, comparing those from Tasso+ capillary blood to those collected via venipuncture. Raw value are shown. GFAP – glial fibrillary acidic protein; NfL – Neurofilament light chain; AB – Amyloid beta.

### Between-Method Correlation and Agreement

Across the 6 biomarkers, between-method correlation (Tasso+ vs. venipuncture) varied widely (Table 3). Aβ40 and Aβ42 demonstrated modest Pearson correlations (0.40–0.47) and were not statistically significant, whereas Spearman correlations were somewhat higher (0.43–0.62) and of marginal significance reflecting rank-order agreement despite substantial differences in absolute concentrations. Aβ42/40 and pTau217 showed poor correlation and negligible concordance, with Lin’s CCC near zero. In contrast, GFAP and NfL showed excellent agreement: Pearson and Spearman correlations exceeded 0.95, and Lin’s CCC approached 0.97–0.98, indicating near-equivalence between Tasso+ and venipuncture.

**Table 3.**
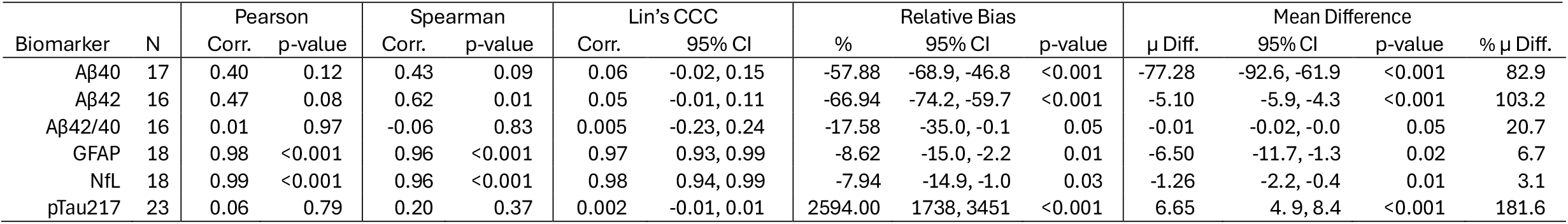
Correlation, agreement, and mean difference of biomarker levels between methods (Tasso+ vs. venipuncture) for the same individuals.

Relative bias analyses showed that Tasso+ systematically underestimated Aβ40 and Aβ42 by 60–70% on average and produced extremely inflated pTau217 values (≈2000%). GFAP and NfL showed small, consistent negative bias (in absolute and relative terms) of only a few percent–yet still statistically different from zero, suggesting minor but consistent underestimation by Tasso+. Pearson & Bland–Altman plots (Figures 4–5) confirmed poor correlation and large disagreement for pTau217 and notable bias for Aβ40 and Aβ42, while GFAP and NfL demonstrated narrow limits of agreement.

**Figure 4.**
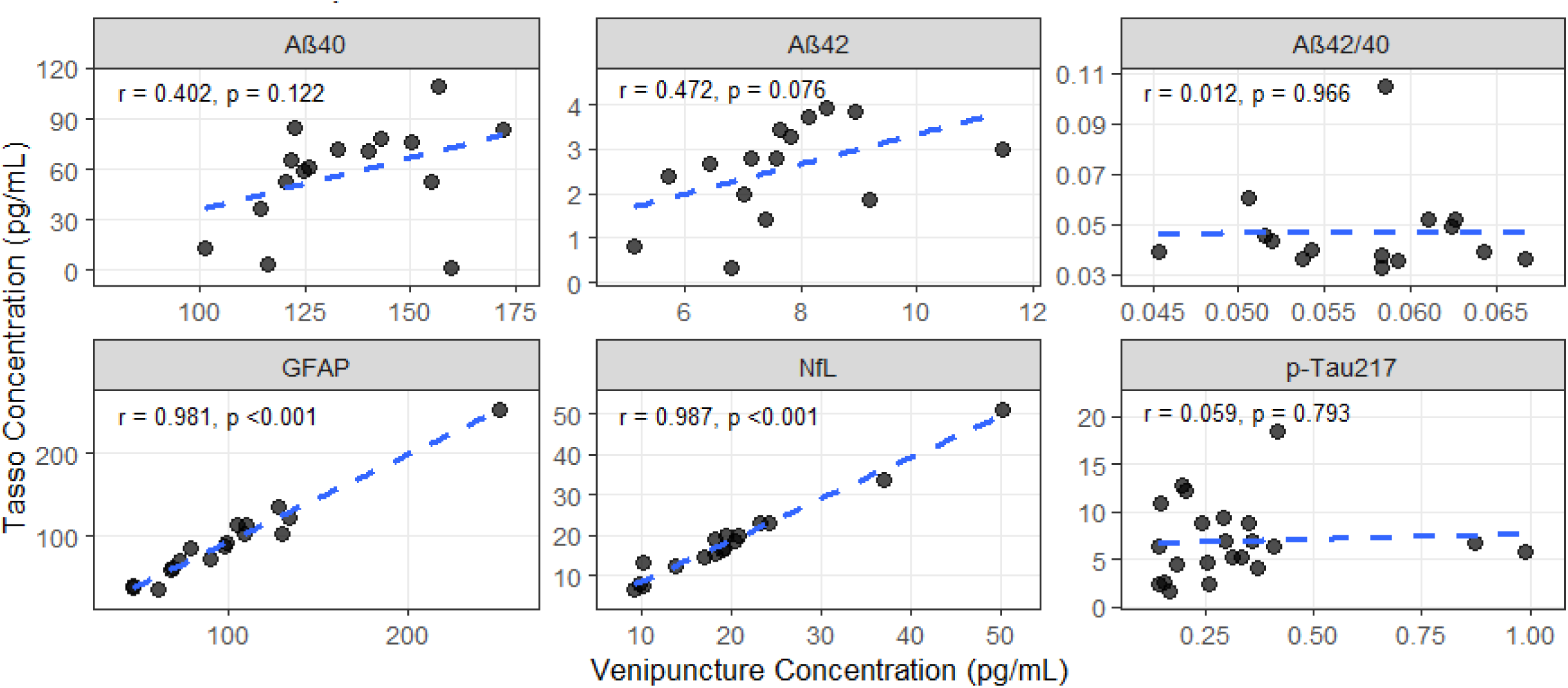
Pearson correlations of ADRD biomarker concentrations in capillary blood from Tasso+ and venous blood samples. GFAP – glial fibrillary acidic protein; NfL – Neurofilament light chain; AB – Amyloid beta.

**Figure 5.**
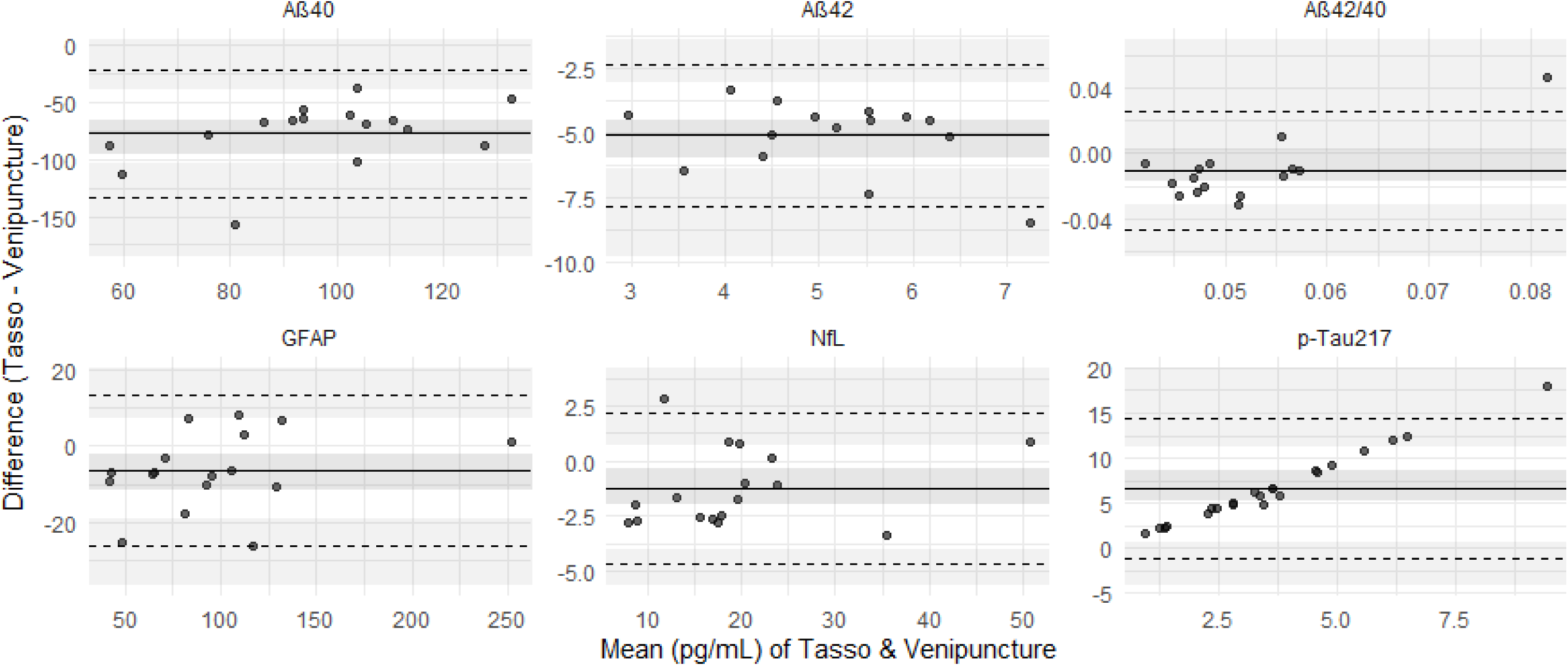
Bland-Altman plots with bootstrapping of the mean of the biomarkers concentrations (pg/mL) from Tasso+ and venipuncture (x-axis) plotted against the difference between the Tasso+ and venipuncture biomarker concentrations. The solid line is the overall mean difference and dashed lines indicate the 95% Confidence Intervals. GFAP – glial fibrillary acidic protein; NfL – Neurofilament light chain; AB – Amyloid beta.

### Within-Method Repeatability

Within-method repeatability was evaluated using duplicate assay runs (Table S5). Venipuncture specimens showed excellent repeatability with low variation across duplicates. For Tasso+, duplicate runs were obtained for a subset of participants for each biomarker (e.g., 8 participants for Aβ40 and GFAP, 6 for Aβ42, 20 for NfL, and 8 for pTau217), whereas nearly all venipuncture samples were run in duplicate. Tasso+ duplicates showed greater variability overall, particularly for Aβ40 and pTau217. For GFAP and NfL, Tasso+ repeatability was strong, with CVs similar to venipuncture. Increased variability in Tasso+ measurements likely reflects pre-analytic factors such as inconsistent volume and hemolysis rather than analytical imprecision alone.

### Predictors of Between-Method Percent Difference

Few participant or collection characteristics were strongly associated with percent mean difference between Tasso+ and venipuncture (Figure 6; Table S6). For most biomarkers, demographic variables were not strongly associated with the Tasso–venipuncture percent difference. For NfL, higher age and evidence of hemolysis in the Tasso+ sample were associated with larger positive percent differences. Hemolysis in Tasso+ was also associated with larger positive percent differences for Aβ40 and Aβ42. For pTau217 and Aβ42/40, some Tasso+ experience variables (e.g., confidence, comfort or willingness) showed associations with percent mean difference, albeit with wide confidence intervals for pTau217. Measures of Tasso+ sample volume, processing lag, and time since last meal were generally not predictive of the differences in biomarkers by collection method, although point estimates were often in the expected direction (e.g., slightly smaller negative bias with larger volumes or shorter processing delays).

**Figure 6.**
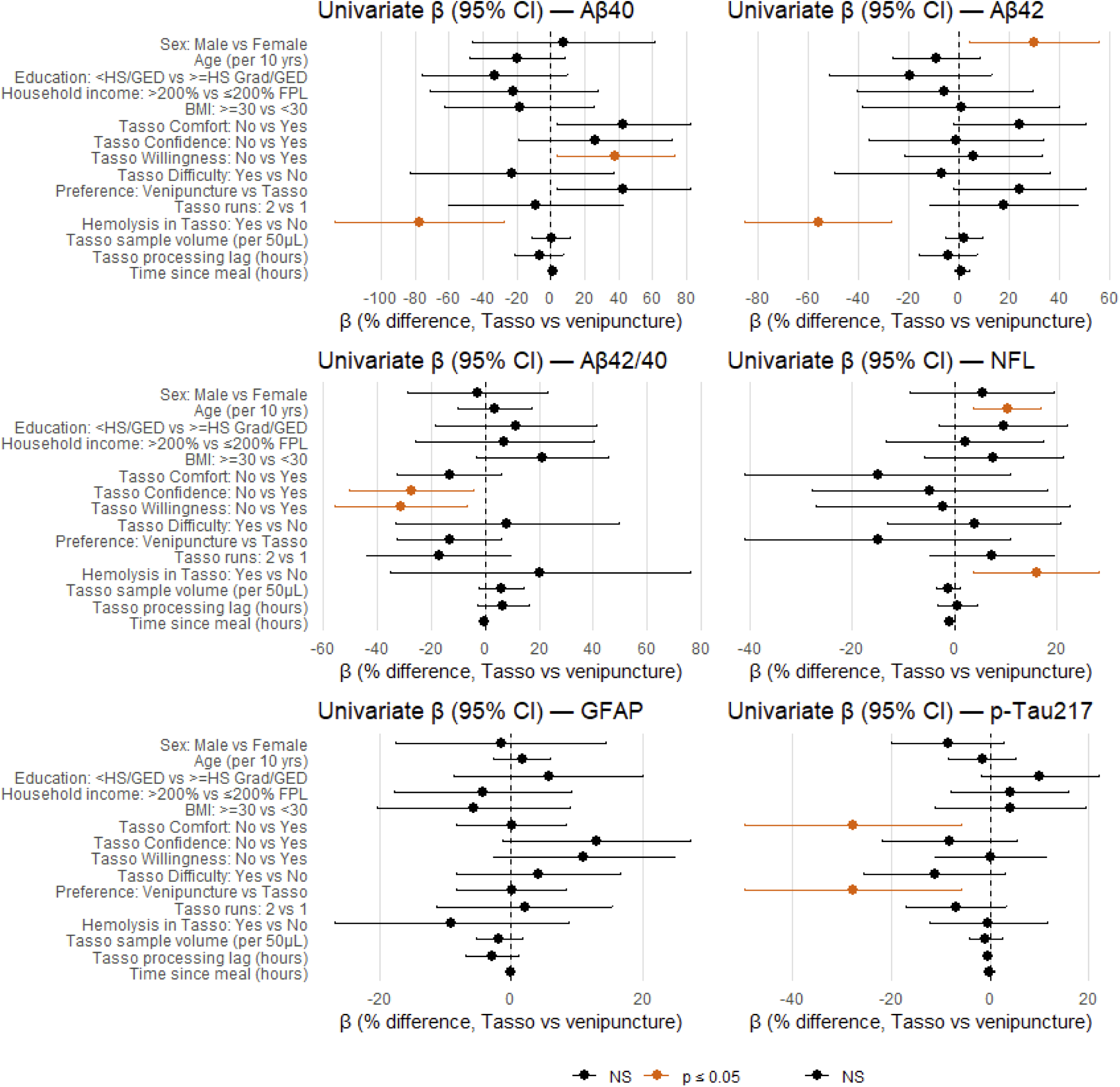
Forest plots depicting beta coefficients (95%CI) of factors associated with the percent mean difference of biomarkers measured via Tasso+ compared with venipuncture (Tasso concentration – Venipuncture concentration). All results are unadjusted; all p-values are unadjusted for multiple comparisons. GFAP – glial fibrillary acidic protein; NfL – Neurofilament light chain; AB – Amyloid beta.

## Discussion

This pilot study successfully recruited a small, but demographically heterogenous sample of adults, reflective of underrepresented populations in Alzheimer’s Disease research in terms of race, ethnicity, educational level, and socio-economic status. This study is among the first to examine the feasibility of using a self-collection blood device, Tasso+, for AD research as a way of increasing representation. Overall, the study found that while most participants had positive experiences self-collecting blood using Tasso+, the majority favored visiting a clinic for venipuncture versus self-collection at home and mailing a blood sample to a lab. Remote capillary blood self-collection with 24-hour delay (to simulate mailing) prior to processing demonstrated high concordance and correlation for GFAP and NfL when compared with venipuncture blood processed immediately; but poor agreement and correlation with core AD biomarkers (AB40, AB42, pTau217).

Participants’ positive experience with using Tasso+, including it being less painful than a venipuncture, aligns with what other studies have documented.^10,14,26^ Yet, among our study sample it was not enough to offset other concerns regarding self-collection. Venipuncture was more often preferred – especially among Black, Hispanic, and lower educated adults in study sample. While Tasso+ may decrease barriers of access to a clinic – a factor influencing low enrollment of underrepresented populations – our study found concerns with mailing blood and the complexity of self-collecting blood overpowered the improved accessibility. Considering the legacy of unethical treatment among racial and ethnic minorities in medical research, this finding is not all that surprising.^2,27^ Future studies seeking to offer self-collection of blood at home among underrepresented populations will need to overcome mistrust in health research and ensure research materials and strategies are culturally sensitive and community informed.^1,28^ For example, the instructional materials provided by Tasso+ will need substantial improvements for comprehension among those with lower education, and materials that adequately address fears around shipping blood from one’s home to a lab should be prioritized and developed with community input.^1,28^ Nonetheless, given people’s positive experience and willingness to use Tasso again in the future, self-collection methods for blood-based AD biomarkers could reach more people.

Capillary blood collection using the Tasso+ device was successful in providing analyzable samples for at least one assay in 82% of attempts, suggesting its feasibility for in-home, self-collection. We found remote capillary blood self-collection with 24-hour shipping prior to processing at a lab is currently a likely suitable method for large-scale studies involving plasma GFAP and NfL, (correlation >0.98, concordance >0.97 when compared to venous blood); yet, we found remote capillary self-blood collection not currently a suitable method for Aβ40, Aβ42, and pTau217. Our findings complement a recent study examining correlation and agreement between proteomics measured via Tasso+ with 24-hour and 48-hour lag prior to processing compared with immediate processing of venipuncture.^13^ This study found a wide range of Pearson correlations and only a minority of proteins had a high correlation and low variation in Tasso samples with venipuncture.^13^

We were not surprised to find low to moderate correlation and agreement between methods for Aβ40 and Aβ42. Several studies have investigated the impact of delayed centrifugation on AD blood biomarker measurements, demonstrating Aβ40 and Aβ42 levels start decreasing after 1-hour and can result in 50% reduction after 24-hour processing delay in both plasma and serum samples.^29,30^ Furthermore, comparison between venous draw versus venous blood spots found high correlation among all AD biomarkers except for Aβ42.^31^ GFAP and NfL have been found to be more stable and degrade less following delayed centrifugation, which we confirmed with the high concordance between methods.^29^ We found pTau217 was severely overestimated by Tasso+, with poor correlation and agreement compared with venipuncture. While delayed processing has not been studied as widely on pTau, pTau217 has been found to deviate from baseline as centrifuge delay increases.^32^ plasma pTau181 was found to notably increase with a 24-, 48- and 72-hour processing delay, but remained stable in serum.^30^ Future method development would benefit from examining pTau217 agreement between remote collection and venipuncture of serum - Tasso+ offers a serum tube attachment. Furthermore, future studies may consider prioritizing specific AD-biomarkers(s) best-suited for the blood derivative (plasma or serum) collected.

The univariate analyses did not reveal a single dominant factor explaining between-method differences; rather, they suggest that method disagreement for Aβ40, Aβ42, and pTau217 reflects a combination of systematic under-recovery in capillary samples and modest contributions from participant-level and pre-analytic factors, whereas GFAP and NfL show minimal systematic bias and limited effect modification. While there was no evidence of longer processing delays being associated with differences in biomarker concentrations, this is likely explained by most Tasso+ samples being processed between 19 and 26 hours post collection, with only one sample exceeding 2 times the 24-hour target. A limitation of the study was our inability to test different post-collection temperatures prior to processing the Tasso+ blood. There is evidence that refrigeration (4°C) for up to 24 hours prior to processing minimizes interference of blood cell lysis and protein degradation.^29^ We have successfully mailed insulated shippers to SHOW participants with cold packs for stool samples^33^ and believe this would be easy to implement to examine how Tasso+ sample shipped at 4°C affects AD biomarkers. Finally, hemolysis was a significant predictor in method disagreement for Aβ40, Aβ42, and NfL, and is known to significantly deteriorate sample quality.^29,34^ It will be important for future studies using Tasso+ to ensure excellent pipetting skills off the mini tubes used with Tasso+.

Overall, venipuncture at a clinic remained the preferred method over Tasso+ self-collected blood at home. However, people’s positive experience with Tasso+ and willingness to use it again in the future indicate it remains an important method to consider for reaching more people for AD research. Correlation and agreement between collection methods were only high for GFAP and NfL. If p-tau, Aβ405, Aβ42, or other markers are of interest, additional specimen collection and handling criteria should be further tested and considered, in particular for maintaining ideal specimen temperatures prior to processing and for reducing hemolysis.

## Supporting information

Survey

## Data Availability

Limited data produced in the present study are available upon reasonable request to the authors

## Acknowledgements

The authors would like to thank the participants for completing the pilot and providing their feedback.

## References

1. Weiner MW, Veitch DP, Miller MJ, et al. Increasing participant diversity in AD research: Plans for digital screening, blood testing, and a community-engaged approach in the Alzheimer’s Disease Neuroimaging Initiative 4. Alzheimers Dement. 2023;19(1):307–317. doi:10.1002/ALZ.12797

2. Babulal GM, Quiroz YT, Albensi BC, et al. Perspectives on ethnic and racial disparities in Alzheimer’s disease and related dementias: Update and areas of immediate need. Alzheimers Dement. 2019;15(2):292–312. doi:10.1016/J.JALZ.2018.09.009

3. Rahman M, White EM, Mills C, Thomas KS, Jutkowitz E. Rural-urban differences in diagnostic incidence and prevalence of Alzheimer’s disease and related dementias. Alzheimers Dement. 2021;17(7):1213–1230. doi:10.1002/ALZ.12285

4. Chen C, Zissimopoulos JM. Racial and ethnic differences in trends in dementia prevalence and risk factors in the United States. Alzheimer’s & Dementia : Translational Research & Clinical Interventions. 2018;4:510. doi:10.1016/J.TRCI.2018.08.009

5. Ashford MT, Raman R, Miller G, et al. Screening and enrollment of underrepresented ethnocultural and educational populations in the Alzheimer’s Disease Neuroimaging Initiative (ADNI). Alzheimers Dement. 2022;18(12):2603–2613. doi:10.1002/ALZ.12640

6. Alawode DOT, Heslegrave AJ, Ashton NJ, et al. Transitioning from cerebrospinal fluid to blood tests to facilitate diagnosis and disease monitoring in Alzheimer’s disease. J Intern Med. 2021;290(3):583–601. doi:10.1111/JOIM.13332

7. Zetterberg H, Schott JM. Blood biomarkers for Alzheimer’s disease and related disorders. Acta Neurol Scand. 2022;146(1):51–55. doi:10.1111/ANE.13628

8. Li D, Mielke MM. An Update on Blood-Based Markers of Alzheimer’s Disease Using the SiMoA Platform. Neurol Ther. 2019;8(Suppl 2):73–82. doi:10.1007/S40120-019-00164-5

9. Zvěřová M. Alzheimer’s disease and blood-based biomarkers - potential contexts of use. Neuropsychiatr Dis Treat. 2018;14:1877–1882. doi:10.2147/NDT.S172285

10. Wickremsinhe E, Fantana A, Berthier E, et al. Standard Venipuncture vs a Capillary Blood Collection Device for the Prospective Determination of Abnormal Liver Chemistry. J Appl Lab Med. 2023;8(3):535–550. doi:10.1093/JALM/JFAC127

11. Brandsma J, Chenoweth JG, Gregory MK, et al. Assessing the use of a micro-sampling device for measuring blood protein levels in healthy subjects and COVID-19 patients. PLoS One. 2022;17(8). doi:10.1371/JOURNAL.PONE.0272572

12. Hameed A, Ferruzzi MG, Kay CD, Williams DK, Rahbar E, Morris AJ. Comparison of the capillary and venous blood plasma lipidomes: validation of self-collected blood for plasma lipidomics. J Lipid Res. 2025;66(3):100755. doi:10.1016/J.JLR.2025.100755

13. El-Sabawi B, Huang S, Tanriverdi K, et al. Capillary blood self-collection for high-throughput proteomics. Proteomics. 2024;24(16):2300607. doi:10.1002/PMIC.202300607;WGROUP:STRING:PUBLICATION

14. Collier BB, Brandon WC, Chappell MR, et al. Comparing capillary blood collection technologies: assessing patient experience, device performance, & clinical accuracy. Bioanalysis. Published online November 1, 2025. doi:10.1080/17576180.2025.2580284;JOURNAL:JOURNAL:IBIO20;WGROUP:STRING:PUBLICATION

15. Huibregtse ME, Sweeney SH, Stephens MR, et al. Association Between Serum Neurofilament Light and Glial Fibrillary Acidic Protein Levels and Head Impact Burden in Women’s Collegiate Water Polo. J Neurotrauma. 2023;40(11-12):1130–1143. doi:10.1089/NEU.2022.0300

16. Planche V, Bouteloup V, Pellegrin I, et al. Validity and Performance of Blood Biomarkers for Alzheimer Disease to Predict Dementia Risk in a Large Clinic-Based Cohort. Neurology. 2023;100(5):E473–E484. doi:10.1212/WNL.0000000000201479

17. Malecki KMC, Nikodemova M, Schultz AA, et al. The Survey of the Health of Wisconsin (SHOW) Program: An Infrastructure for Advancing Population Health. Front Public Health. 2022;10:818777. doi:10.3389/FPUBH.2022.818777/BIBTEX

18. U.S. Census Bureau. Census urban and rural classification and urban area criteria. Commerce UDo, editor. 2020. Accessed December 6, 2025. https://www.census.gov/programs-surveys/geography/guidance/geo-areas/urban-rural.html

19. Harris PA, Taylor R, Minor BL, et al. The REDCap consortium: Building an international community of software platform partners. J Biomed Inform. 2019;95. doi:10.1016/j.jbi.2019.103208

20. Nasreddine ZS, Phillips NA, Bédirian V, et al. The Montreal Cognitive Assessment, MoCA: a brief screening tool for mild cognitive impairment. J Am Geriatr Soc. 2005;53(4):695–699. doi:10.1111/J.1532-5415.2005.53221.X

21. Therriault J, Servaes S, Tissot C, et al. Equivalence of plasma p-tau217 with cerebrospinal fluid in the diagnosis of Alzheimer’s disease. Alzheimers Dement. 2023;19(11):4967–4977. doi:10.1002/ALZ.13026

22. Bayoumy S, Verberk IMW, den Dulk B, et al. Clinical and analytical comparison of six Simoa assays for plasma P-tau isoforms P-tau181, P-tau217, and P-tau231. Alzheimer’s Research & Therapy. 2021;13(1). doi:10.1186/s13195-021-00939-9

23. Department of Health and Human Services. Annual Update of the HHS Poverty Guidelines. Federal Register, 89(11), 2961–2963. January 17, 2024. Accessed December 6, 2025. https://www.federalregister.gov/documents/2024/01/17/2024-00796/annual-update-of-the-hhs-poverty-guidelines

24. Centers for Disease Control and Prevention. Adult BMI Categories. CDC. March 19, 2024. Accessed December 6, 2025. https://www.cdc.gov/bmi/adult-calculator/bmi-categories.html

25. Kind AJH, Buckingham WR. Making Neighborhood-Disadvantage Metrics Accessible — The Neighborhood Atlas. New England Journal of Medicine. 2018;378(26):2456–2458. doi:10.1056/NEJMP1802313/SUPPL_FILE/NEJMP1802313_DISCLOSURES.PDF

26. Schröder D, Hafke A, Hummers E, et al. Comparison of laboratory results and pain perception in self-sampled capillary blood versus venous blood sampling: a systematic review and meta-analysis. Clin Biochem. 2025;138:110965. doi:10.1016/J.CLINBIOCHEM.2025.110965

27. Lincoln KD, Chow T, Gaines BF, Fitzgerald T. Fundamental causes of barriers to participation in Alzheimer’s clinical research among African Americans. Ethn Health. 2021;26(4):585–599. doi:10.1080/13557858.2018.1539222

28. Mindt MR, Okonkwo O, Weiner MW, et al. Improving generalizability and study design of Alzheimer’s disease cohort studies in the United States by including under-represented populations. Alzheimers Dement. 2023;19(4):1549–1557. doi:10.1002/ALZ.12823

29. Verberk IMW, Misdorp EO, Koelewijn J, et al. Characterization of pre-analytical sample handling effects on a panel of Alzheimer’s disease–related blood-based biomarkers: Results from the Standardization of Alzheimer’s Blood Biomarkers (SABB) working group. Alzheimer’s & Dementia. 2021;18(8):1484. doi:10.1002/ALZ.12510

30. Panikkar D, Vivek S, Crimmins E, Faul J, Langa KM, Thyagarajan B. Pre-Analytical Variables Influencing Stability of Blood-Based Biomarkers of Neuropathology. J Alzheimers Dis. 2023;95(2):735. doi:10.3233/JAD-230384

31. Huber H, Blennow K, Zetterberg H, et al. Biomarkers of Alzheimer’s disease and neurodegeneration in dried blood spots—A new collection method for remote settings. Alzheimer’s and Dementia. 2024;20(4):2340–2352. doi:10.1002/ALZ.13697

32. Figdore DJ, Schuder BJ, Ashrafzadeh-Kian S, Gronquist T, Bornhorst JA, Algeciras-Schimnich A. Differences in Alzheimer’s disease blood biomarker stability: Implications for the use of tau/amyloid ratios. Alzheimer’s & Dementia. 2025;21(4):e70173. doi:10.1002/ALZ.70173

33. Schultz AA, Malecki KM, Holzhausen EA, et al. The Population-based Microbiome Research Core: a longitudinal infrastructure for assessment of household microbiome and human health research. doi:10.1101/2021.11.22.21266369

34. Ozga MR, Bittner T, Batrla R, Karl J. Preanalytical sample handling recommendations for Alzheimer’s disease plasma biomarkers. doi:10.1016/j.dadm.2019.02.002

