## Supplementary tables and figures for "Feasibility and validity of using self-collected capillary blood using Tasso+ for measuring Alzheimer’s Disease plasma-based biomarkers among underrepresented populations"

**SUPPLEMENTARY MATERIALS**

| **Table S1.** Collection method preferences (Tasso vs. venipuncture) and feedback from free response items grouped by themes | |
| --- | --- |
| **What was the most confusing step of using the Tasso+ device?** | **n** |
| Making sure it "clicked" for it to work (wasn't clear) | 4 |
| Waiting for the blood and knowing if it's working | 2 |
| Putting the device in the correct spot | 2 |
| Activating the heat pad | 1 |
| Need to hang arm straight (wasn't clear) | 1 |
| Closing up the tube. "Twist" was not clear in instructions. | 1 |
| 5-minute limit should be specified better | 1 |
| Mirror seemed optional, but it isn't | 1 |
| Seeing the video with visual impairment | 1 |
| Not knowing how the blood should mix | 1 |
| **What was the most difficult part of using the Tasso+ device?** | **n** |
| Waiting for the blood to come out, knowing if its working | 5 |
| Not knowing how hard to push button, that it should click | 4 |
| Getting the tube off the Tasso and getting Tasso off | 3 |
| Getting started; lots of steps to follow | 2 |
| Lining the tube up to see the lines | 1 |
| Anticipating pain but none came | 1 |
| **Why Tasso was preferred over venous draw** | **n** |
| **Easier / more convenient.** | **8** |
| *Easier* |  |
| *less time commitment* |  |
| *more convenient at home* |  |
| *don't have to leave house & go to hospital* |  |
| *"I live in the middle of nowhere"* |  |
| *better for regular, frequent check-ups* |  |
| **Would be good method if there is an outbreak.** | **1** |
| **Why venous draw was preferred over Tasso** | **n** |
| **Easier.** | **7** |
| *Don't want to have to mail it* |  |
| *Don't want to have to do the blood collection myself* |  |
| **Tasso+ was difficult.** | **4** |
| *Don't think Tasso+ works well for heaver set folks* |  |
| *Had difficulty getting enough blood* |  |
| *People may have difficulty reading instructions; may not have*  *internet for video* |  |
| *Too difficult with neuropathy / arthritis* |  |
| **Concerns about Tasso+ sample.** | **2** |
| *Worried sample may get lost in mail; quality of sample may not be*  *as good as venous draw* |  |
| *Concerned quality of sample may not be as good as venous draw* |  |
| **Like to see people.** | **2** |

| **Table S2.** Blood collection status by method (n=28) | | |
| --- | --- | --- |
|  | n | % |
| **Was venipuncture blood collected?** |  |  |
| Yes | 26 | 93 |
| No, but attempted | 2 | 7 |
| **Was Tasso blood collected?** |  |  |
| Yes, by participant | 25 | 89 |
| Yes, by phlebotomist^a^ | 2 | 7 |
| No, but attempted | 1 | 4 |
| **Number of Tasso collection attempts** |  |  |
| one attempt | 24 | 86 |
| two attempts | 3 | 10 |
| three attempts | 1 | 4 |
| **^a^** n=1 visually impaired; n=1 with neuropathy & was difficult to push button in | | |

| **Table S3.** Number of samples with hemolysis and lab results by sample collection method. | | | |
| --- | --- | --- | --- |
| **Hemolysis status** | | **Tasso**  (n=27)  n | **Venipuncture** (n=26)  n |
| None | | 17 | 25 |
| Slight | | 3 | 0 |
| Moderate | | 2 | 1 |
| Full | | 5 | 0 |
| *No sample* | | *1* | *2* |
| **Useable sample** | |  |  |
| Aβ40 | | 17 | 26 |
| Aβ42 | | 16 | 26 |
| GFAP | | 18 | 26 |
| NfL | | 18 | 26 |
| pTau217 | | 23 | 26 |
| **Singlet or duplicate analysis** | |  |  |
| Aβ40 | Single | 9 | 0 |
|  | Double | 8 | 26 |
| Aβ42 | Single | 10 | 0 |
|  | Double | 6 | 26 |
| GFAP | Single | 10 | 0 |
|  | Double | 8 | 26 |
| NfL | Single | 3 | 0 |
|  | Double | 20 | 26 |
| pTau217 | Single | 9 | 1 |
|  | Double | 8 | 25 |

| **Table S4.** Hemolysis status by total plasma volume collected via Tasso device. | | | | | | |
| --- | --- | --- | --- | --- | --- | --- |
| Tasso plasma volume (uL) | Total | | Hemolysis^a^ | | Un-useable Sample | |
|  | n | % | n | % | N | % |
| 0 | 1 | 4 | n/a | n/a | n/a | n/a |
| 75 | 2 | 7 | 1 | 50 | 2 | 100 |
| 100 | 6 | 21 | 1 | 17 | 1 | 17 |
| 150 | 3 | 11 | 0 | 0 | 0 | 0 |
| 200 | 7 | 25 | 2 | 29 | 2 | 29 |
| 250 | 5 | 18 | 2 | 40 | 1 | 20 |
| 300 | 2 | 7 | 1 | 50 | 1 | 50 |
| 350 | 0 | 0 | n/a | n/a | n/a | n/a |
| 400 | 1 | 4 | 0 | 0 | 0 | 0 |
| 450 | 0 | 0 | n/a | n/a | n/a | n/a |
| 500 | 1 | 4 | 0 | 0 | 0 | 0 |

^a^Only includes moderate to full hemolysis (n=7); did not include slight hemolysis (n=3)

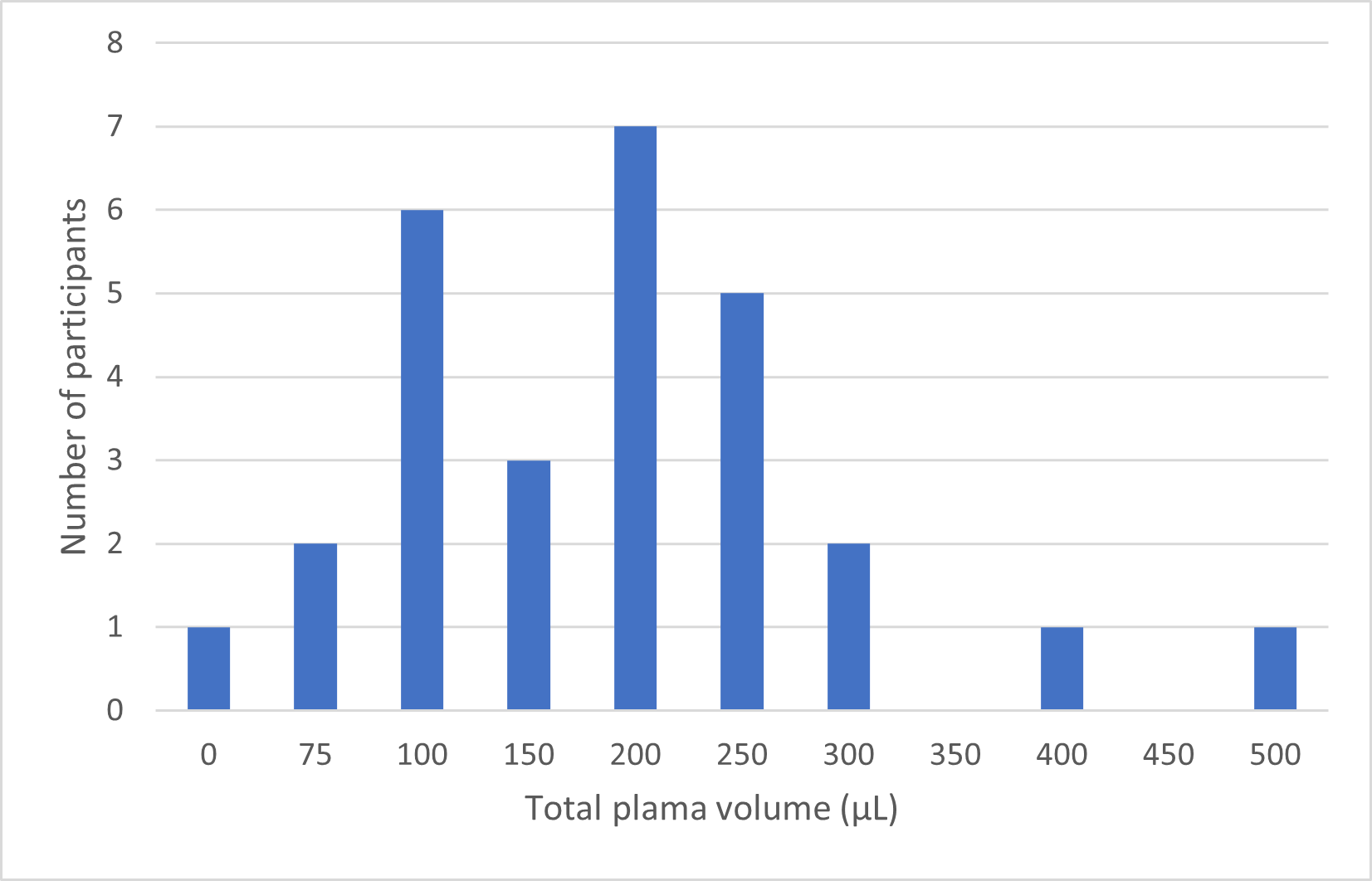

**Figure S1.** Distribution of total plasma volume collected by Tasso+ (n=27)

| 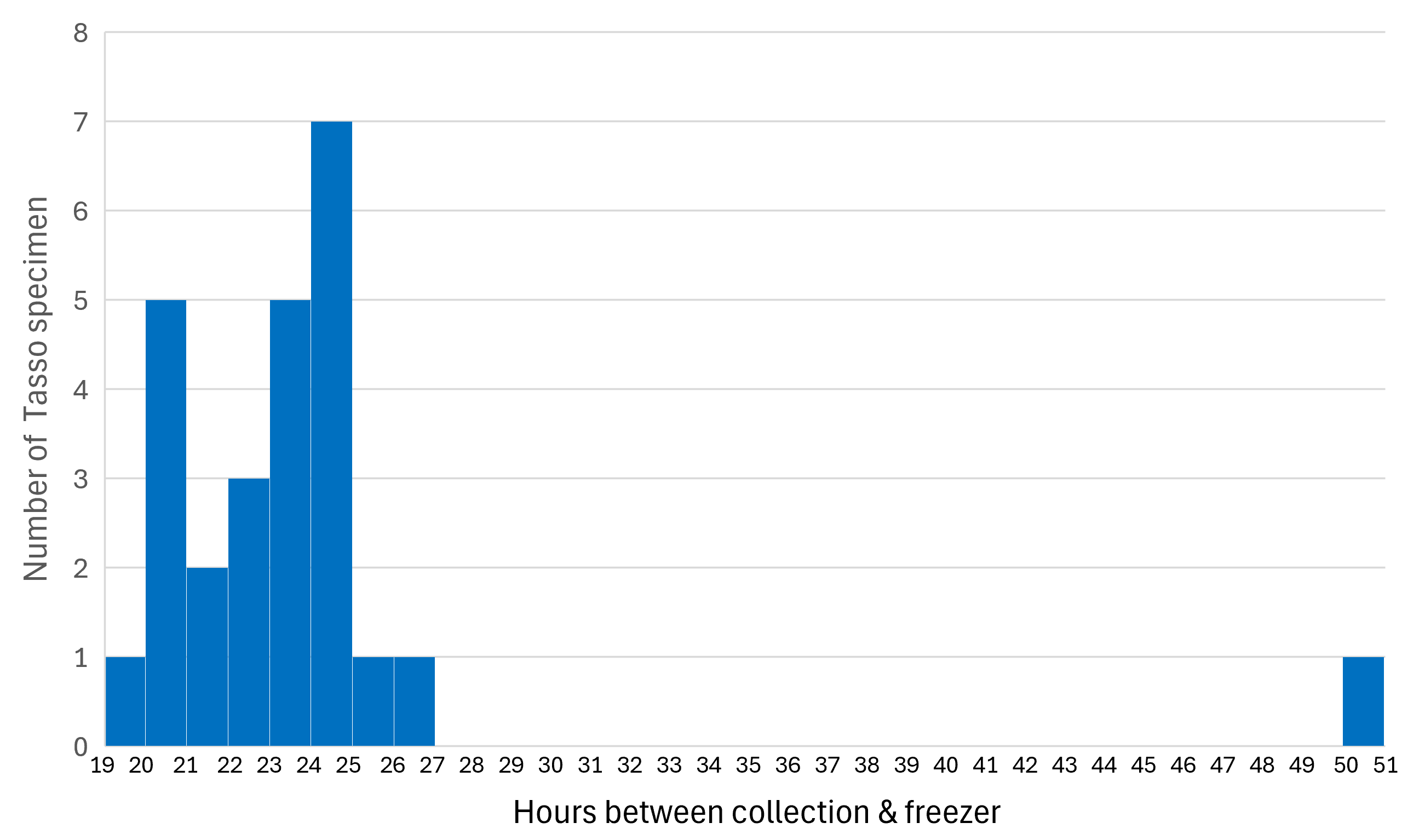  **Figure S2.** Distribution of time lag between collection and freezing plasma volume collected by Tasso+ (n=27)  **Table S5**. Comparison of summary statistics and between-participant standard deviation (SD) and coefficient of variation (CV) by the number of runs (singlet, duplicate, or either) that was conducted for each biomarker. | | | | | | | | | | | | | |
| --- | --- | --- | --- | --- | --- | --- | --- | --- | --- | --- | --- | --- | --- |
|  | No. of runs | Tasso | | | | | | Venipuncture | | | | | |
| Biomarker |  | N | Min | Max | Mean | SD | CV | N | Min | Max | Mean | SD | CV |
| Aβ40 | 1 | 9 | 1.99 | 84.87 | 56.77 | 24.90 | 43.86 | 0 |  |  |  |  |  |
|  | 2 | 8 | 3.10 | 109.15 | 58.82 | 35.56 | 60.46 | 26 | 101.49 | 205.65 | 139.44 | 25.25 | 18.11 |
|  | 1 or 2 | 17 | 1.99 | 109.15 | 57.73 | 29.40 | 50.93 | n/a |  |  |  |  |  |
| Aβ42 | 1 | 10 | 0.32 | 3.82 | 2.23 | 1.12 | 50.30 | 0 |  |  |  |  |  |
|  | 2 | 6 | 1.98 | 3.91 | 3.04 | 0.77 | 25.30 | 26 | 5.13 | 11.47 | 7.93 | 1.56 | 19.69 |
|  | 1 or 2 | 16 | 0.32 | 3.91 | 2.53 | 1.06 | 41.68 | n/a |  |  |  |  |  |
| GFAP | 1 | 10 | 35.68 | 135.32 | 75.75 | 33.49 | 44.21 | 0 |  |  |  |  |  |
|  | 2 | 8 | 62.40 | 252.71 | 112.16 | 61.17 | 54.54 | 26 | 38.28 | 251.63 | 98.32 | 51.54 | 52.42 |
|  | 1 or 2 | 18 | 35.68 | 252.71 | 91.94 | 49.81 | 54.18 | n/a |  |  |  |  |  |
| NfL | 1 | 10 | 6.50 | 23.24 | 14.40 | 6.12 | 42.49 | 0 |  |  |  |  |  |
|  | 2 | 8 | 13.00 | 51.05 | 23.47 | 12.80 | 54.55 | 26 | 6.13 | 50.21 | 19.00 | 10.14 | 53.36 |
|  | 1 or 2 | 18 | 6.50 | 51.05 | 18.43 | 10.43 | 56.59 | n/a |  |  |  |  |  |
| pTau217 | 1 | 3 | 2.60 | 8.79 | 5.91 | 3.12 | 52.79 | 1 | 0.21 | 0.21 | n/a | n/a | n/a |
|  | 2 | 20 | 1.71 | 18.42 | 7.14 | 4.05 | 56.73 | 25 | 0.14 | 0.99 | 0.34 | 0.22 | 63.34 |
|  | 1 or 2 | 23 | 1.71 | 18.42 | 6.98 | 3.90 | 55.93 | 26 | 0.14 | 0.99 | 0.34 | 0.21 | 63.52 |

| **Table S6**. Univariate factors associated with greater percent mean difference between biomarkers measured via Tasso+ compared with venipuncture. All results are unadjusted; all p-values are unadjusted for multiple comparisons. | | | | | | | | | | | | |
| --- | --- | --- | --- | --- | --- | --- | --- | --- | --- | --- | --- | --- |
| **Biomarker** | **Predictor** | **Comparison** | | **n** | | **β** | | **CI Lower** | | **CI Upper** | | **p-value** |
| **Aβ40** | Sex | Female vs Male | | 16 | | - 7.54 | | -61.52 | | 46.44 | | 0.79 |
|  | Age | *per 10-year increase* | | 16 | | -20.06 | | -48.18 | | 8.05 | | 0.18 |
|  | Education | >=HS Grad/GED vs <HS/GED | | 16 | | 32.79 | | -10.21 | | 75.78 | | 0.16 |
|  | Household income | >200% vs <=200% FPL | | 15 | | -21.91 | | -71.44 | | 27.62 | | 0.40 |
|  | BMI | >=30 vs <30 | | 16 | | -18.44 | | -62.49 | | 25.61 | | 0.43 |
|  | **Tasso Comfort** | **No vs Yes** | | **16** | | **42.87** | | **3.63** | | **82.12** | | **0.05** |
|  | Tasso Confidence | No vs Yes | | 16 | | 26.13 | | -19.33 | | 71.60 | | 0.28 |
|  | **Tasso Willingness** | **No vs Yes** | | **16** | | **38.36** | | **3.60** | | **73.12** | | **0.05** |
|  | Tasso Difficulty | Yes vs No | | 16 | | -22.91 | | -82.90 | | 37.09 | | 0.47 |
|  | **Preference** | **Venipuncture vs Tasso** | | **16** | | **42.87** | | **3.63** | | **82.12** | | **0.05** |
|  | Tasso runs | 2 vs 1 | | 16 | | - 8.90 | | -60.38 | | 42.59 | | 0.74 |
|  | **Hemolysis in Tasso** | **Yes vs No** | | **16** | | **-77.41** | | **-127.40** | | **-27.42** | | **0.01** |
|  | Tasso sample volume | *per 50 µL increase* | | 16 | | 0.00 | | - 0.22 | | 0.23 | | 0.97 |
|  | Tasso processing lag | *per 1 hour increase* | | 16 | | - 6.64 | | -21.10 | | 7.82 | | 0.38 |
|  | Time since meal | *per 1 hour increase* | | 16 | | 1.36 | | - 0.54 | | 3.26 | | 0.18 |
| **Aβ42** | **Sex** | Female vs Male | | 15 | | -30.15 | | -56.01 | | - 4.29 | | **0.04** |
|  | Age | *per 10-year increase* | | 15 | | - 8.77 | | -26.08 | | 8.53 | | 0.34 |
|  | Education | >=HS Grad/GED vs <HS/GED | | 15 | | 19.11 | | -13.14 | | 51.36 | | 0.27 |
|  | Household income | >200% vs <=200% FPL | | 14 | | - 5.54 | | -40.60 | | 29.51 | | 0.76 |
|  | BMI | >=30 vs <30 | | 15 | | 0.97 | | -38.26 | | 40.20 | | 0.96 |
|  | Tasso Comfort | No vs Yes | | 15 | | 24.27 | | - 1.90 | | 50.44 | | 0.09 |
|  | Tasso Confidence | No vs Yes | | 15 | | - 0.84 | | -35.56 | | 33.88 | | 0.96 |
|  | Tasso Willingness | No vs Yes | | 15 | | 5.89 | | -21.30 | | 33.08 | | 0.68 |
|  | Tasso Difficulty | Yes vs No | | 15 | | - 6.54 | | -49.34 | | 36.26 | | 0.77 |
|  | Preference | Venipuncture vs Tasso | | 15 | | 24.27 | | - 1.90 | | 50.44 | | 0.09 |
|  | Tasso runs | 2 vs 1 | | 15 | | 18.10 | | -11.26 | | 47.47 | | 0.25 |
|  | **Hemolysis in Tasso** | **Yes vs No** | | 15 | | -55.79 | | -85.00 | | -26.58 | | **0.002** |
|  | Tasso sample volume | *per 50 µL increase* | | 15 | | 0.04 | | - 0.11 | | 0.19 | | 0.58 |
|  | Tasso processing lag | *per 1 hour increase* | | 15 | | - 4.14 | | -15.71 | | 7.43 | | 0.50 |
|  | Time since meal | *per 1 hour increase* | | 15 | | 1.33 | | - 1.42 | | 4.08 | | 0.36 |
| **Aβ42/40** | Sex | Female vs Male | | 15 | | 2.88 | | -23.02 | | 28.78 | | 0.83 |
|  | Age | | *per 10-year increase* | | 15 | | 3.48 | | -10.42 | | 17.38 | 0.63 |
|  | Education | | >=HS Grad/GED vs <HS/GED | | 15 | | -11.47 | | -41.26 | | 18.33 | 0.46 |
|  | Household income | | >200% vs <=200% FPL | | 14 | | 7.05 | | -26.07 | | 40.18 | 0.68 |
|  | BMI | | >=30 vs <30 | | 15 | | 21.31 | | - 3.13 | | 45.75 | 0.11 |
|  | Tasso Comfort | | No vs Yes | | 15 | | -13.33 | | -32.84 | | 6.18 | 0.20 |
|  | **Tasso Confidence** | | **No vs Yes** | | **15** | | **-27.46** | | **-50.45** | | **- 4.47** | **0.04** |
|  | **Tasso Willingness** | | **No vs Yes** | | **15** | | **-31.20** | | **-55.71** | | **- 6.69** | **0.03** |
|  | Tasso Difficulty | | Yes vs No | | 15 | | 8.16 | | -33.45 | | 49.78 | 0.71 |
|  | Preference | | Venipuncture vs Tasso | | 15 | | -13.33 | | -32.84 | | 6.18 | 0.20 |
|  | Tasso runs | | 2 vs 1 | | 15 | | -17.25 | | -43.82 | | 9.32 | 0.23 |
|  | Hemolysis in Tasso | | Yes vs No | | 15 | | 20.32 | | -35.20 | | 75.84 | 0.49 |
|  | Tasso sample volume | | *per 50 µL increase* | | 15 | | 0.12 | | - 0.05 | | 0.28 | 0.18 |
|  | Tasso processing lag | | *per 1 hour increase* | | 15 | | 6.64 | | - 2.91 | | 16.20 | 0.20 |
|  | Time since meal | | *per 1 hour increase* | | 15 | | - 0.21 | | - 1.56 | | 1.15 | 0.77 |

| **Table S5 (Continued)** | | | | | | | |
| --- | --- | --- | --- | --- | --- | --- | --- |
| **Biomarker** | **Predictor** | **Comparison** | **n** | **β** | **CI Lower** | **CI Upper** | **p-value** |
| **GFAP** | Sex | Female vs Male | 17 | 1.52 | -14.46 | 17.49 | 0.85 |
|  | Age | *per 10-year increase* | 17 | 1.74 | - 2.62 | 6.10 | 0.45 |
|  | Education | >=HS Grad/GED vs <HS/GED | 17 | - 5.72 | -20.17 | 8.74 | 0.45 |
|  | Household income | >200% vs <=200% FPL | 16 | - 4.27 | -17.74 | 9.19 | 0.54 |
|  | BMI | >=30 vs <30 | 17 | - 5.59 | -20.28 | 9.10 | 0.47 |
|  | Tasso Comfort | No vs Yes | 17 | 0.10 | - 8.30 | 8.50 | 0.98 |
|  | Tasso Confidence | No vs Yes | 17 | 13.02 | - 1.21 | 27.25 | 0.09 |
|  | Tasso Willingness | No vs Yes | 17 | 11.11 | - 2.64 | 24.85 | 0.13 |
|  | Tasso Difficulty | Yes vs No | 17 | 4.19 | - 8.26 | 16.64 | 0.52 |
|  | Preference | Venipuncture vs Tasso | 17 | 0.10 | - 8.30 | 8.50 | 0.98 |
|  | Tasso runs | 2 vs 1 | 17 | 2.12 | -11.27 | 15.50 | 0.76 |
|  | Hemolysis in Tasso | Yes vs No | 17 | - 9.01 | -26.75 | 8.74 | 0.34 |
|  | Tasso sample volume | *per 50 µL increase* | 17 | - 0.03 | - 0.10 | 0.03 | 0.34 |
|  | Tasso processing lag | *per 1 hour increase* | 17 | - 2.81 | - 6.78 | 1.15 | 0.18 |
|  | Time since meal | *per 1 hour increase* | 17 | - 0.05 | - 0.54 | 0.44 | 0.84 |
| **NFLight** | Sex | Female vs Male | 17 | - 5.51 | -19.57 | 8.56 | 0.45 |
|  | **Age** | ***per 10-year increase*** | **17** | **10.30** | **3.74** | **16.86** | **0.01** |
|  | Education | >=HS Grad/GED vs <HS/GED | 17 | - 9.52 | -22.04 | 2.99 | 0.16 |
|  | Household income | >200% vs <=200% FPL | 16 | 2.08 | -13.28 | 17.43 | 0.79 |
|  | BMI | >=30 vs <30 | 17 | 7.70 | - 5.88 | 21.27 | 0.28 |
|  | Tasso Comfort | No vs Yes | 17 | -14.97 | -40.91 | 10.98 | 0.28 |
|  | Tasso Confidence | No vs Yes | 17 | - 4.75 | -27.70 | 18.20 | 0.69 |
|  | Tasso Willingness | No vs Yes | 17 | - 2.23 | -26.92 | 22.46 | 0.86 |
|  | Tasso Difficulty | Yes vs No | 17 | 3.89 | -13.07 | 20.85 | 0.66 |
|  | Preference | Venipuncture vs Tasso | 17 | -14.97 | -40.91 | 10.98 | 0.28 |
|  | Tasso runs | 2 vs 1 | 17 | 7.31 | - 4.90 | 19.53 | 0.26 |
|  | **Hemolysis in Tasso** | **Yes vs No** | **17** | **16.08** | **3.82** | **28.34** | **0.02** |
|  | Tasso sample volume | *per 50 µL increase* | 17 | - 0.02 | - 0.07 | 0.02 | 0.34 |
|  | Tasso processing lag | *per 1 hour increase* | 17 | 0.57 | - 3.25 | 4.40 | 0.77 |
|  | Time since meal | *per 1 hour increase* | 17 | - 0.94 | - 1.97 | 0.09 | 0.10 |
| **pTau217** | Sex | Female vs Male | 21 | 8.57 | - 2.73 | 19.88 | 0.15 |
|  | Age | *per 10-year increase* | 22 | - 1.57 | - 8.48 | 5.34 | 0.66 |
|  | Education | >=HS Grad/GED vs <HS/GED | 22 | -10.17 | -22.22 | 1.89 | 0.11 |
|  | Household income | >200% vs <=200% FPL | 21 | 4.05 | - 7.97 | 16.07 | 0.52 |
|  | BMI | >=30 vs <30 | 22 | 4.14 | -11.15 | 19.42 | 0.60 |
|  | **Tasso Comfort** | **No vs Yes** | **22** | **-27.82** | **-49.74** | **- 5.90** | **0.02** |
|  | Tasso Confidence | No vs Yes | 22 | - 8.19 | -21.97 | 5.58 | 0.26 |
|  | Tasso Willingness | No vs Yes | 22 | 0.14 | -11.15 | 11.43 | 0.98 |
|  | Tasso Difficulty | Yes vs No | 22 | -11.20 | -25.58 | 3.18 | 0.14 |
|  | **Preference** | **Venipuncture vs Tasso** | **22** | **-27.82** | **-49.74** | **- 5.90** | **0.02** |
|  | Tasso runs | 2 vs 1 | 22 | - 6.86 | -16.95 | 3.23 | 0.20 |
|  | Hemolysis in Tasso | Yes vs No | 22 | - 0.30 | -12.14 | 11.54 | 0.96 |
|  | Tasso sample volume | *per 50 µL increase* | 22 | - 0.02 | - 0.08 | 0.05 | 0.61 |
|  | Tasso processing lag | *per 1 hour increase* | 22 | - 0.39 | - 1.18 | 0.41 | 0.35 |
|  | Time since meal | *per 1 hour increase* | 22 | - 0.03 | - 0.88 | 0.81 | 0.94 |

| **Table S7**. Among those with replicate runs, the within-participant concentration (conc.) means and ranges, and standard deviation (SD), and coefficient of variation (CV) by biomarker and collection method. | | | | | | | | | | | |
| --- | --- | --- | --- | --- | --- | --- | --- | --- | --- | --- | --- |
| Biomarker | Collection | N | Min conc. | Max conc. | Mean conc. | Replicate SD (min) | Replicate SD (max) | Replicate SD (mean) | Replicate CV% (min) | Replicate CV% (max) | Replicate CV% (mean) |
| Abeta40 | Tasso | 8 | 3.10 | 109.15 | 58.82 | 0.008 | 3.516 | 1.370 | 0.000 | 0.104 | 0.035 |
|  | Venipuncture | 26 | 101.49 | 205.65 | 139.44 | 0.251 | 11.519 | 3.886 | 0.002 | 0.097 | 0.028 |
| Abeta42 | Tasso | 6 | 1.98 | 3.910 | 3.04 | 0.017 | 0.168 | 0.103 | 0.004 | 0.063 | 0.038 |
|  | Venipuncture | 26 | 5.13 | 11.47 | 7.93 | 0.009 | 0.618 | 0.203 | 0.001 | 0.113 | 0.027 |
| GFAP | Tasso | 8 | 62.40 | 252.71 | 112.16 | 0.045 | 3.337 | 1.615 | 0.001 | 0.044 | 0.017 |
|  | Venipuncture | 26 | 38.28 | 251.63 | 98.32 | 0.116 | 5.521 | 2.451 | 0.002 | 0.123 | 0.030 |
| NFLight | Tasso | 8 | 13.00 | 51.05 | 23.47 | 0.119 | 1.292 | 0.697 | 0.007 | 0.060 | 0.033 |
|  | Venipuncture | 26 | 6.13 | 50.21 | 19.00 | 0.003 | 1.769 | 0.588 | 0.000 | 0.104 | 0.037 |
| pTau217 | Tasso | 20 | 1.71 | 18.42 | 7.14 | 0.000 | 1.004 | 0.297 | 0.000 | 0.140 | 0.040 |
|  | Venipuncture | 25 | 0.14 | 0.990 | 0.34 | 0.001 | 0.031 | 0.010 | 0.003 | 0.171 | 0.034 |
