## Supplementary material for "Feasibility and validity of using self-collected capillary blood using Tasso+ for measuring Alzheimer’s Disease plasma-based biomarkers among underrepresented populations": Survey

### Tasso+ Blood Collection

Record ID ______________________

**When was the last time you had anything to eat or drink other than water?**
Date: ______
Time: ______

**Which arm would you prefer for the self-collected blood sample (I will perform the blood draw on the other arm)?**
- ◯ Right
- ◯ Left

**Next, you will perform a self-collected blood sample. One of the main purposes of this study is to determine whether people feel comfortable collecting their own blood at home.**

**One goal of this study is to determine what kind of instructions are helpful. Paper instructions will be sent with the collection device in the mail. The instructions will also have a link where you can watch a short video explaining how to complete the self-collection. I am going to ask you to first read the paper instructions, and then watch the video instructions. Afterwards, I will ask you a couple questions before we continue.**

Play Tasso+ Video

(participant reads and watches instructional info)

**Which directions did you find most helpful? The paper, video, or both?**
- ◯ Paper
- ◯ Video
- ◯ Both
- ◯ Neither

**Do you have any questions or concerns before you begin?**
(record questions, and answer questions before beginning)

__________

**Before you begin, do you plan to follow along with the video (you can pause it as you go)?Or do you prefer to follow the paper instructions?**

(help the participant get set up as needed)

Tasso+ Blood Collection Occurs

What instructions did the participant reference during Tasso+ collection:
- ◯ Video
- ◯ Paper
- ◯ Video and Paper
- ◯ Neither Video or Paper

Was blood collected using Tasso+?
- ◯ Yes- by participant
- ◯ Yes- by phleb
- ◯ No- but attempted
- ◯ No- refused

Date and time of capillary self collection: ______________________

Explain any issues that arose during the Tasso+ collection: __________________________________________

Record any comments made by the participant: __________________________________________

### Tasso+ Feedback Survey

Record ID __________________________________

Tasso Feedback Survey

**How useful were the Tasso instructions in guiding you through the collection process?**

- ◯ Not useful
- ◯ Slightly useful
- ◯ Moderately useful
- ◯ Very useful

**What was the most confusing step of using the Tasso+ device?** __________________________________

**How easy or difficult was it to use the Tasso+ device?**

- ◯ Very easy
- ◯ Somewhat easy
- ◯ Neither easy nor difficult
- ◯ Somewhat difficult
- ◯ Very difficult

**What was the most difficult part of using the Tasso+ device?** __________________________________________

**What did you feel when using the Tasso+ device?**

- ◯ Comfort / no pain
- ◯ Slight discomfort / slight pain
- ◯ Moderate discomfort / moderate pain
- ◯ Severe discomfort / severe pain

**How willing are you to use a Tasso+ device in the future?**

- ◯ Very willing
- ◯ Somewhat willing
- ◯ Undecided
- ◯ Somewhat unwilling
- ◯ Not willing

**How confident are you in your ability to follow the instructions and use the Tasso+ on your own in the future?**

- ◯ Very confident
- ◯ Somewhat confident
- ◯ Neither confident nor unconfident
- ◯ Somewhat unconfident
- ◯ Not confident

### Venous Blood Draw & Feedback Survey

Record ID ______________________________

Was venous blood collected?
- ◯ Yes
- ◯ No- but attempted
- ◯ No- refused

Date and time of blood draw ______________________________

Right or left arm?
- ◯ Right arm
- ◯ Left arm

**When compared to the normal blood draw I just performed, was the Tasso+ more painful, less painful, or about the same?**
- ◯ Tasso+ was more painful
- ◯ Tasso+ was about the same
- ◯ Tasso+ was less painful

**If given the choice between completing a self-collected blood sample at home using Tasso+ (and mailing to a lab) vs. going to a clinic for a normal blood draw, which would you choose?**
- ◯ Tasso+
- ◯ Normal blood draw

**Why do you prefer the Tasso+?** ______________________________

**Why do you prefer the normal blood draw?** ______________________________

**Is there any feedback you would like to provide on your experience using Tasso+ that you have not provided yet?**

__________
